# Predicting the risks for stroke, cardiovascular disease, and peripheral vascular disease among people with type 2 diabetes with artificial intelligence models: a systematic review and meta-analysis

**DOI:** 10.1101/2024.08.13.24311939

**Authors:** Aqsha Nur, Sydney Tjandra, Defin Yumnanisha, Arnold Keane, Adang Bachtiar

## Abstract

**Objectives:** This systematic review and meta-analysis aim to explore the performance of machine learning algorithms in predicting the risk of macrovascular complications among individuals with T2DM, specifically, the predictive capabilities of AI models in forecasting stroke, CVD, and PVD in LMICs.

**Design:** Systematic review and meta-analysis of studies reporting on AI prediction models for macrovascular complications in T2DM patients.

**Setting:** The review included studies conducted in various healthcare settings, primarily from LMICs, upper-middle-income countries (UMICs), and high-income countries (HICs).

**Participants:** 46 studies were included, with a total of 184 AI models. Participants were diverse in age, sex, and geographical locations, reflecting a broad range of healthcare settings.

**Interventions:** The intervention analyzed was the application of AI models, including machine learning algorithms, to predict macrovascular complications such as stroke, CVD, and PVD.

**Primary and Secondary Outcome Measures:** The primary outcome was the predictive performance of AI models, measured by the area under the receiver operating characteristic curve (AUROC). Secondary outcomes included subgroup analyses based on predictor types and an assessment of AI model applicability in low-resource settings.

**Results:** Twelve included studies yielded 184 AI models with an overall AUROC of 0.753 (95%CI: 0.74-0.766; I2=99.99%; p<0.001). For 80 models of cardiovascular outcomes, an AUROC of 0.741 (95%CI: 0.721-0.76; I2=99.78%; p<0.001) was obtained. Meanwhile, 25 models of peripheral vascular disease and 38 models of cerebrovascular diseases obtained AUROCs of 0.794 (95%CI: 0.758-0.831; I2=97.23%; p<0.001) and 0.77 (95%CI: 0.743-0.797; I2=99.73%; p<0.001) respectively. Subgroup analysis revealed that models with lab-only predictors were superior to those with mixed or no-lab predictors. This signalled the lack of AI capability for history-taking and physical examination data alone, primarily available in low-resource settings.

**Conclusions:** Artificial intelligence is promising in predicting diabetes complications. Nevertheless, future studies should explore accessible features in low-resource settings and employ external validation to ensure the robustness of the prediction models.

**Article Summary:** *Strengths and limitations of this study:*

- Inclusion of studies from both health-related and computer science databases (such as IEEE Xplore) ensured a comprehensive assessment of AI models for predicting diabetes complications.
- The study analyzed a wide range of models from various countries with different income levels, enhancing the generalizability of the findings.
- Detailed subgroup analyses provided insights into the impact of predictor types (lab vs. non-lab) and machine learning algorithms on model performance.
- High heterogeneity across studies, stemming from variations in populations, data sources, and algorithms, was observed, reflecting a common issue in AI model performance meta-analyses.
- A significant limitation was the lack of external validation in most included studies, which raises concerns about the generalizability and applicability of the AI models in diverse clinical settings.

*Key Messages:*

- What is already known on this topic

- Artificial intelligence (AI) holds great potential for diabetes care.
- Previous meta-analyses have shown its promise in diabetes predictions, but none has been done for diabetes complication predictions.
- What this study adds

- AI model performance aggregates provided promising results.
- Subgroup analyses exposed characteristics facilitating prediction performances, namely gradient-boosting algorithms, lab predictors, cross- validation, and detailed missing data.
- How this study might affect research, practice, or policy

- Albeit promising, ethical open-source models enabling multiple external validations and interdisciplinary collaboration are vital before broader implementation.

## Introduction

At least 500 million people were estimated to live with diabetes in 2021, of which 96% were type 2 diabetes mellitus (T2DM).^1^ Diabetes complications, such as stroke, cardiovascular diseases (CVD), and peripheral vascular diseases (PVD), increase the 5-year mortality, particularly for people living in low-and middle-income countries (LMICs).^2^ According to World Health Organization (WHO) estimates, 75% of CVD deaths occur in LMICs.^3^ As the global burden continues to rise, there is an urgent need for precise and early risk stratification methods to enable timely preventive measures for T2DM complications.^4^ In this context, the use of artificial intelligence (AI) and machine learning (ML) models has garnered significant interest for their potential to enhance predictive accuracy in the management of T2DM complications.^5^ ^6^ These technologies promise to transform traditional healthcare approaches by leveraging vast amounts of data to uncover complex patterns and relationships that may not be readily apparent through conventional statistical methods.^7^

Previous systematic reviews have primarily focused on the potential of AI in various aspects of diabetes care, particularly in predicting the onset of diabetes itself. For instance, recent meta-analyses have demonstrated the utility of AI in forecasting diabetes-related outcomes, yet none have comprehensively addressed the prediction of macrovascular complications associated explicitly with T2DM.^8–10^ This gap highlights the necessity of a focused investigation into how AI can be harnessed to predict complications like stroke, CVD, and PVD in patients already diagnosed with T2DM, including its deployment in low-resource settings.^11^

This systematic review and meta-analysis aim to expdiabalore the performance of machine learning algorithms in predicting the risk of macrovascular complications among individuals with T2DM, specifically, the predictive capabilities of AI models in forecasting stroke, CVD, and PVD. Unlike earlier studies that may have concentrated on a single type of complication or generalized diabetes prediction, this review delves into a broader spectrum. Moreover, it provides an in-depth analysis of subgroup performances, comparing models with various predictor types, including lab-only and mixed predictors, and examining the implications of these differences. This review also highlights the challenges and limitations associated with current AI models, particularly their applicability in low-resource settings. By focusing on models that utilize widely available data and require minimal specialized input, the findings might guide future research or policy-making for AI tools that can be deployed effectively in regions with limited healthcare infrastructure.

## Materials and Methods

### Search Strategy

This review was systematically developed, conducted, and reported following Preferred Reporting Items for Systematic Review and Meta-Analysis (PRISMA) checklist during writing our report as presented in the Supplementary Material 1.^12^ Our protocol has been registered at The International Prospective Register of Systematic Reviews (PROSPERO) under the reference ID CRD42023489167.

We searched six databases (Scopus, PubMed, Embase, Wiley Online Library, IEEE Explore, and Google Scholar) and hand-searched for articles published between January 1, 2000, and November 30, 2023. Keywords employed were “type 2 diabetes”, “artificial intelligence,” “prediction,” “complication,” “stroke,” “cardiovascular disease,” and “peripheral vascular disease,” as well as their MeSH terms and subsets combined with Boolean operators (see Supplementary Material 2). Search results were exported and deduplicated to Rayyan (www.rayyan.ai).

### Eligibility Criteria

Each article was screened for the following PICOT inclusion criteria (see Supplementary Material 3) by at least two members independently (AN, ST, RH, SW):^13^ (1) subjects are adults aged 18 years old or above with type 2 diabetes mellitus, (2) intervention was the development and implementation of artificial intelligence, including but not limited to machine learning and deep learning, as opposed to classical statistical models, (3) outcome included prediction performances for stroke, cardiovascular disease, or peripheral vascular disease, (4) diagnostic or prognostic studies with a cohort or case-control design capable of exhibiting temporality, (5) used any actual medical dataset, and (6) published in English. We excluded studies that (1) had mixed populations with type 1 and/or prediabetes patients, (2) mainly explained theoretical models not tested on human subjects, (3) involved drugs as the intervention, (4) were reviews, framework developments, conference abstracts, proposals, editorials, commentaries, and qualitative studies, and (5) had irretrievable full-text. After titles and abstracts were screened on Rayyan, full-text screening was conducted to reconfirm eligibility. Discrepancies were resolved through consensus.

### Data Extraction

A data extraction instrument was developed to tribulate several characteristics and details from all included studies, namely (1) author and year, (2) country of origin, (3) study design, (4) data source, (5) single or multi-centred, (6) population profile (including number of patients, age, and proportion of males), (7) predictors, (8) whether external validation was employed, (9) AI/ML algorithm, (10) outcome (stroke, cardiovascular disease, or peripheral vascular disease), (11) data period and follow-up, (12) data pre-processing details, and (13) internal validation setup. We also extracted the main outcome, model performance, in metrics such as F-measures, the area under the receiving operating curve (AUROC), c-statistics, sensitivity/recall, specificity, accuracy, and precision/positive predictive value.

### Risk of Bias Assessment

Two members (AN, ST, DY, AK) independently assessed all included studies for risk of bias and applicability using the signalling questions on the Prediction Model Risk of Bias Assessment Tool (PROBAST).^14^ Differences were discussed to reach a consensus.

### Quantitative Data Analysis

Studies reporting AUROCs as model performances were aggregated through a random-effects meta-analysis with MedCalc, visualized with R. When neither the standard error, range, nor the standard deviation was disclosed, we ran the Hanley and McNeil’s approach^15^ with R to approximate the standard error based on the AUROC, sample size, and number of complication cases.^15–17^ To assess publication bias, funnel plots and Egger’s regression were generated with MedCalc. Moreover, outliers, defined as models whose 95% confidence intervals did not overlap with the meta-analysis result, were excluded to generate sensitivity analyses. As substantial heterogeneity remains, subgroup analyses were conducted for outcome types, external validation, algorithms, country income levels, risk of bias, missing data process details, cross-validation, and predictor data.

## Results and Discussion

### Study Characteristics

A total of 2,513 studies were found during the initial search across seven databases (PubMed, Google Scholar, EMBASE, Scopus, IEEE Explore, and Wiley), in addition to hand searching. After removing 512 duplicate records, 2,001 records were screened for their titles and abstracts. Subsequently, 1,895 records were excluded, leaving 106 reports to be retrieved. Studies without available full texts were excluded, resulting in 95 studies being assessed for eligibility. Of these, 49 studies were excluded for various reasons: unsuitable population (17 studies), irrelevant outcome (29 studies), unsuitable study design (2 studies), and text not in English (1 study). Ultimately, 46 studies were included in the systematic review, with 30 included in the quantitative analysis. The selection process is depicted in Figure 1.

**Figure 1:**
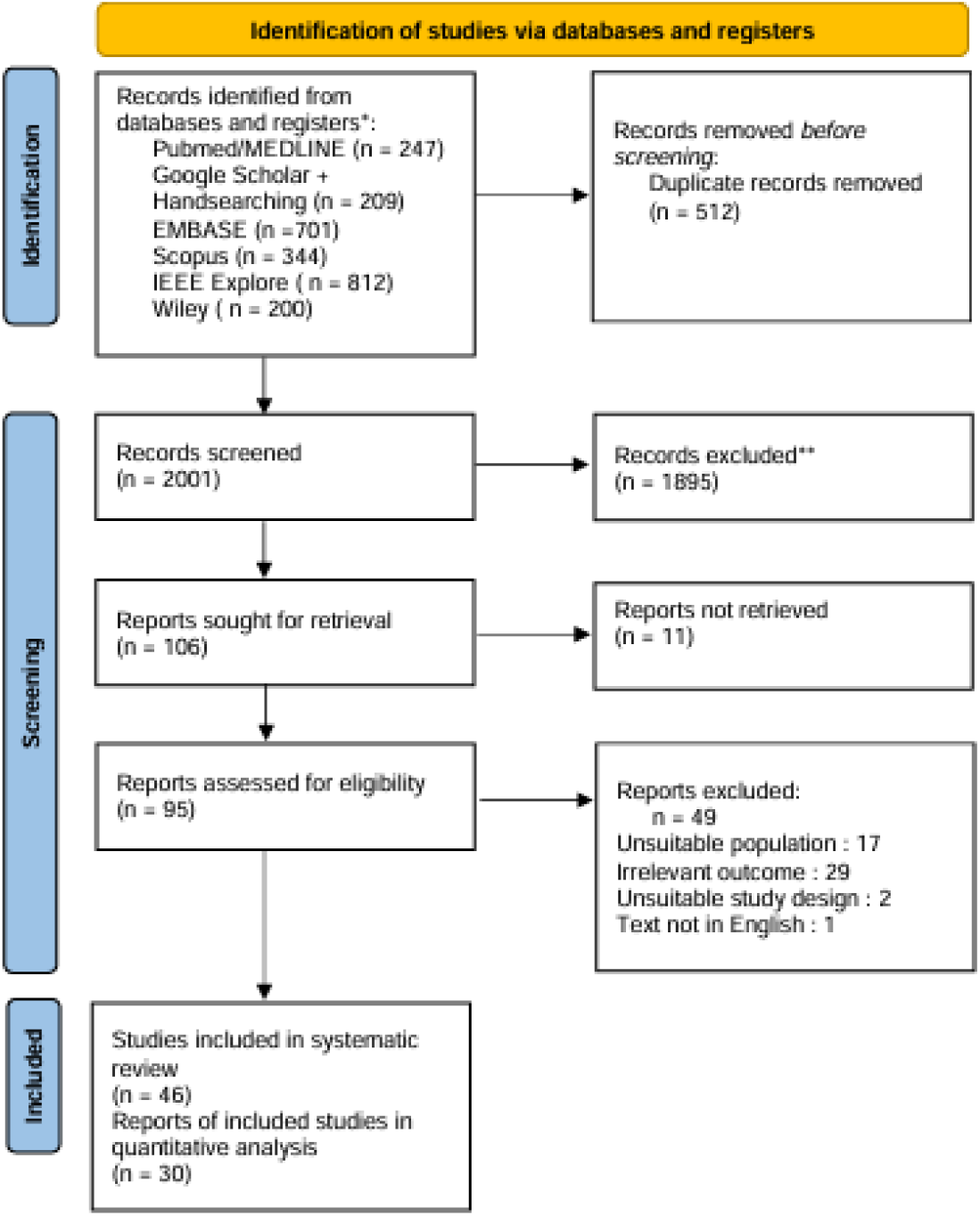
PRISMA flowchart of included studies.

Supplementary Material 4 depicts the overall characteristics of the study, including the participants and outcomes of each study. The systematic review encompasses 46 studies from various countries, which are categorized into three regions based on income levels: low-middle-income countries (LMIC), upper-middle-income countries (UMIC), and high-income countries (HIC). Most data were sourced from hospital medical records in the respective countries, while some datasets were from specific trials or studies. Sample sizes from each study varied from around a hundred to hundreds of thousands, even surpassing a million. The predictors were divided into demographic, clinical, comorbidity, and laboratory groups. The algorithms were classified into several main categories, such as Random Forest, Neural Network, Gradient Boosting, Logistic Regression, Multi-task Learning, Cox-based methods, and others. The outcomes assessed included cardiovascular disease, cerebrovascular disease (stroke), and peripheral vascular disease. Information about technical aspects such as handling missing data, cross-validation, and external validation is also provided.

### Risk of Bias

The overall risk of bias, as summarised in Figure 2, indicates that 78% of the articles are rated as having a high or uncertain risk of bias, with specific distributions of 10 articles rated as low risk, ten as unclear risk, and 27 as high risk. The high risk of bias predominantly originated from the “outcome” domain due to the uncertainty in determining outcomes without knowledge of predictor information. Additionally, the “analysis” domain contributed significantly to the high risk, primarily due to inadequate handling of missing data or improper imputation methods and the low number of participants with the outcome. Nearly all studies (91%) showed no concerns regarding applicability concerns.

**Figure 2:**
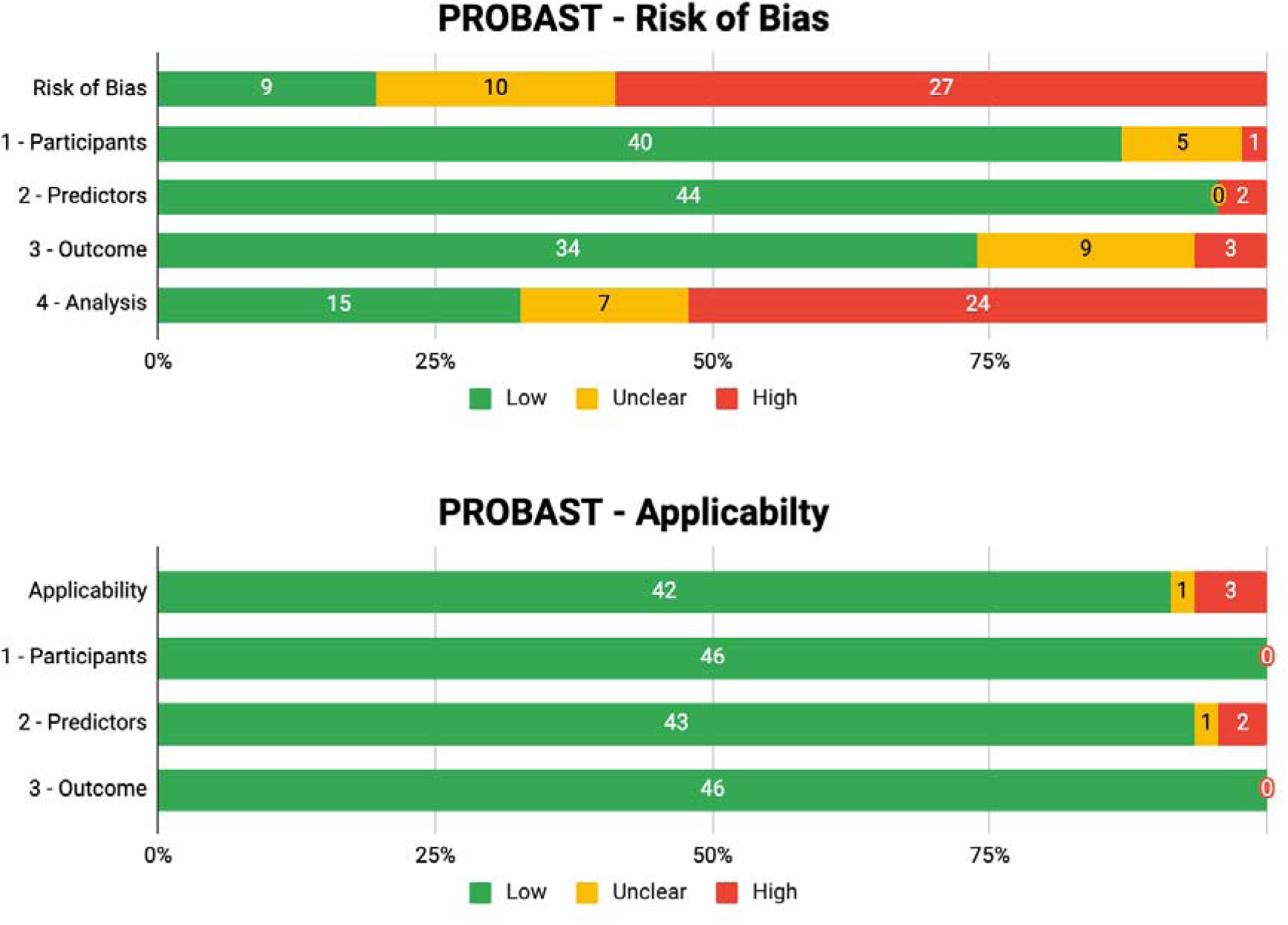
PROBAST summary of included studies

### Meta-Analysis of AUROCs

All 184 models were pooled with a random effects model, obtaining an AUROC of 0.753 (95%CI: 0.74–0.766; I^2^=99.99%; p<0.001), as shown through the forest plot in Figure 3. For 80 models of cardiovascular outcomes, an AUROC of 0.741 (95%CI: 0.721–0.76; I^2^=99.78%; p<0.001) was obtained. Meanwhile, 25 models of peripheral vascular disease and 38 models of cerebrovascular diseases obtained AUROCs of 0.794 (95%CI: 0.758–0.831; I^2^=97.23%; p<0.001) and 0.77 (95%CI: 0.743–0.797; I^2^=99.73%; p<0.001) respectively (Supplementary Materials 6–9). Subgroup analysis results are detailed in Table 1.

**Figure 3:**
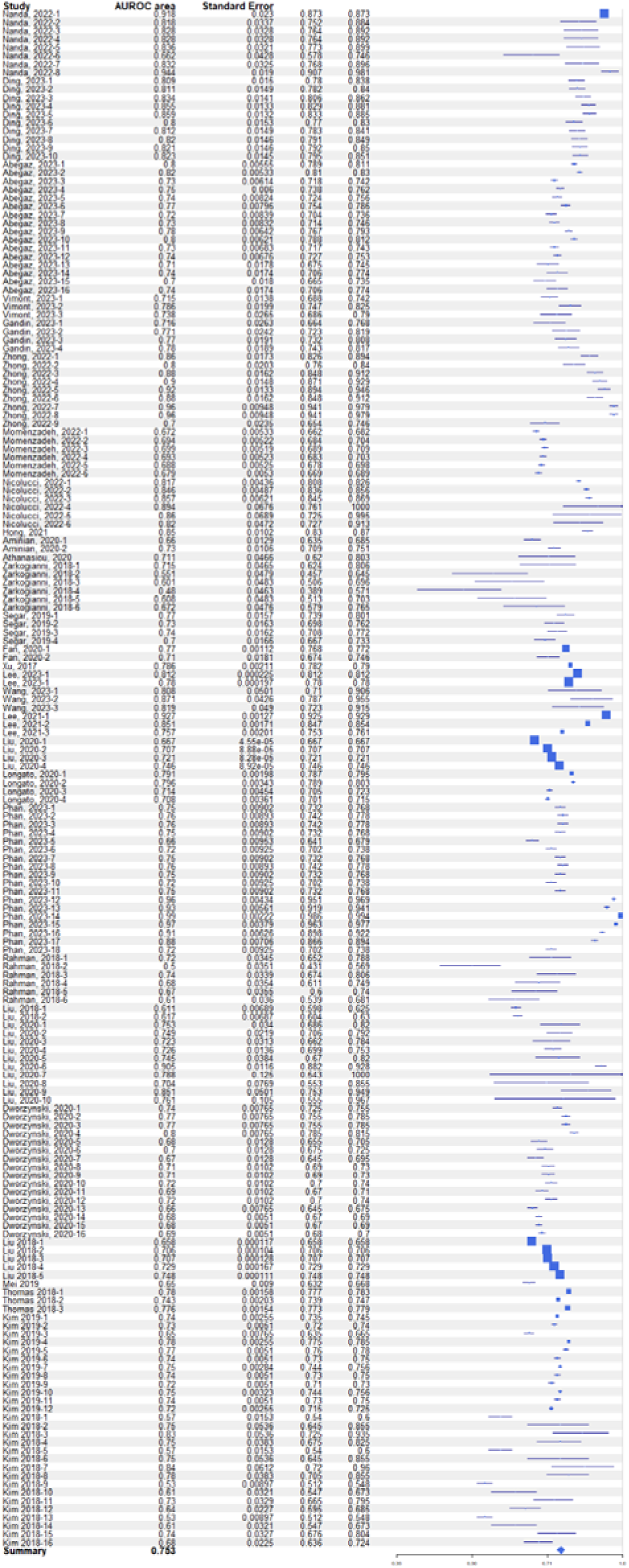
Forest plot of artificial intelligence areas under the operating curve (AUROCs) in predicting diabetes complications.

**Table 1:** Subgroup analyses summary of Area Under the Receiver Operating Characteristics (AUROCs) of machine learning prediction models for diabetes macrovascular complications based on various characteristics.

| Subgroups | No. of prediction models | Random-effects AUROC | Lower 95% CI | Higher 95% CI | Heterogeneity (%) | p-value |
| --- | --- | --- | --- | --- | --- | --- |
| All studies | 184 | 0.753 | 0.74 | 0.766 | 99.99 | < 0.001 |
| **Outcome types** |  |  |  |  |  |  |
| Cardiovascular | 80 | 0.741 | 0.721 | 0.76 | 99.78 | < 0.001 |
| Peripheral vascular/diabetic foot | 25 | 0.794 | 0.758 | 0.831 | 97.23 | < 0.001 |
| Stroke / cerebrovascular | 38 | 0.77 | 0.743 | 0.797 | 99.73 | < 0.001 |
| Mixed | 41 | 0.741 | 0.716 | 0.765 | 100 | < 0.001 |
| **External validation data** |  |  |  |  |  |  |
| Yes | 56 | 0.725 | 0.708 | 0.742 | 97.85 | < 0.001 |
| No | 128 | 0.765 | 0.749 | 0.782 | 99.99 | < 0.001 |
| **Machine learning algorithm** |  |  |  |  |  |  |
| Cox-based | 14 | 0.712 | 0.664 | 0.76 | 98.53 | < 0.001 |
| Gradient boosting | 37 | 0.789 | 0.761 | 0.817 | 99.56 | < 0.001 |
| Logistic regression | 23 | 0.731 | 0.711 | 0.752 | 99.55 | < 0.001 |
| Multi-task learning | 18 | 0.699 | 0.665 | 0.733 | 99.99 | < 0.001 |
| Neural network | 11 | 0.759 | 0.722 | 0.797 | 98.55 | < 0.001 |
| Random forest | 30 | 0.776 | 0.742 | 0.810 | 99.73 | < 0.001 |
| Others | 51 | 0.752 | 0.726 | 0.777 | 99.99 | < 0.001 |
| **Country income** |  |  |  |  |  |  |
| HIC | 134 | 0.737 | 0.723 | 0.751 | 99.99 | < 0.001 |
| LIC/LMIC/UMIC | 50 | 0.8 | 0.774 | 0.825 | 97.31 | < 0.001 |
| **Risk of bias** |  |  |  |  |  |  |
| Low/medium | 110 | 0.780 | 0.765 | 0.794 | 99.76 | < 0.001 |
| High | 74 | 0.711 | 0.691 | 0.731 | 99.99 | < 0.001 |
| **Missing data process detailed** |  |  |  |  |  |  |
| Yes | 114 | 0.775 | 0.76 | 0.79 | 99.66 | < 0.001 |
| No | 70 | 0.717 | 0.696 | 0.738 | 99.99 | < 0.001 |
| **Cross-validation** |  |  |  |  |  |  |
| Yes | 127 | 0.759 | 0.743 | 0.775 | 99.99 | < 0.001 |
| No | 57 | 0.739 | 0.717 | 0.761 | 98.88 | < 0.001 |
| **Predictor data** |  |  |  |  |  |  |
| No lab | 29 | 0.714 | 0.696 | 0.731 | 100 | < 0.001 |
| Lab only | 3 | 0.837 | 0.784 | 0.89 | 0 | < 0.001 |
| Mixed | 152 | 0.759 | 0.745 | 0.774 | 99.98 | < 0.001 |

AUROC, Area Under the Receiver Operating Characteristic; CI, confidence interval; HIC, high-income countries; LIC, low-income countries; LMIC, lower-middle-income countries; UMIC, upper-middle-income countries

### Publication Bias

We observed significant publication bias (Egger’s test p-value = 0.0261). Funnel plots of AUROCs against standard errors are presented in Supplementary Materials 10—14.

### Sensitivity Analyses

We excluded outliers and retrieved 83 models with an overall AUROC of 0.746 (95%CI: 0.742–0.75; I^2^=99.86%; p<0.001). This is comparable to the initial meta-analysis, showing robustness despite outliers. Similarly, outcome and predictor subgroup sensitivity analyses were conducted, with results in Table 2. Most notably, the peripheral vascular disease outcome subgroup retrieved an AUROC of 0.820 (95%CI: 0.798–0.842; p<0.001) with a heterogeneity of I^2^=0%. In the lab-only predictors subgroup, no outliers were identified.

**Table 2:** Sensitivity analyses summary of Area Under the Receiver Operating Characteristics (AUROCs) of machine learning prediction models for diabetes macrovascular complications based on outcomes and predictors.

| Subgroups | No. of prediction models | Random-effects AUROC | Lower 95% CI | Higher 95% CI | Heterogeneity (%) | p-value |
| --- | --- | --- | --- | --- | --- | --- |
| All studies | 83 | 0.746 | 0.742 | 0.75 | 99.86 | < 0.001 |
| **Outcome types** |  |  |  |  |  |  |
| Cardiovascular | 38 | 0.741 | 0.733 | 0.749 | 80.99 | < 0.001 |
| Peripheral vascular/diabetic foot | 15 | 0.820 | 0.798 | 0.842 | 0.00 | < 0.001 |
| Stroke / cerebrovascular | 18 | 0.756 | 0.747 | 0.764 | 92.42 | < 0.001 |
| Mixed | 16 | 0.737 | 0.729 | 0.745 | 99.97 | < 0.001 |
| **External validation data** |  |  |  |  |  |  |
| No lab | 15 | 0.710 | 0.703 | 0.718 | 99.90 | < 0.001 |
| Mixed | 73 | 0.753 | 0.748 | 0.758 | 92.31 | < 0.001 |

## Discussion

### Model Performance

The pooled analysis of 184 models revealed a moderate level of performance, with an overall AUROC of 0.753. The models demonstrated varying performance based on the specific outcome types. The models achieved an AUROC of 0.741 for cardiovascular disease outcomes, while those predicting peripheral vascular disease and cerebrovascular disease had higher AUROCs of 0.794 and 0.77, respectively. Nanda R et al. (2022) generated an RF model with the highest AUC across all models of 0.918 to predict the risk of T2DM people developing diabetic foot ulcers.^18^ These results suggest that while the models are generally robust, their effectiveness can vary depending on the specific type of predicted macrovascular complication.

Heterogeneity among the included studies was notably high, with I^2^ values approaching 100% across most analyses. This substantial heterogeneity underscores the variability in model performance, which could stem from differences in study populations, data sources, predictor variables, and machine learning algorithms used. The high heterogeneity highlights the importance of context-specific factors in model performance and suggests that predictive accuracy may improve when models are tailored to specific populations and settings. The sensitivity analysis further supports the robustness of the findings. By excluding outliers, the overall AUROC was slightly reduced to 0.746, with heterogeneity remaining high (I^2^=99.86%). This consistency indicates that extreme values do not unduly influence the overall conclusions.

Using these ML models for diabetes complication risk prediction might be helpful in considering several limitations in several existing conventional scoring systems. For example, the Framingham risk score, the most established risk assessment for heart disease, was developed for the general population and not specific for T2DM people.^19^ This risk score is designed for the general population and not specifically for people with diabetes. Risk scores developed for general populations may have lower discriminatory ability in individuals with diabetes.^20^ Other researchers have also developed heart disease risk assessment focus using ML models,^21^ with linear and logistic regression, and artificial neural networks (ANN) often used due to their simplicity and good predictive ability.

### Study settings

Most studies (29; 63%) in our review came from research in HICs, followed by UMICs (9; 19.6%) and LMICs (8; 17.4%). India accounts for the majority of studies from the LMICs, China dominates the UMICs, and the United States leads in the HICs. This might demonstrate that a country’s income level influences the number of artificial intelligence research and publications. A bibliometric study by Jimma (2023) mapped the publication of artificial intelligence in hea Interestingly, the United States and China are included in the top nine countries, with the United States ranking first (41.84%) and China second (14.70%). Our study also identified that the most productive and prominent institutions funding AI research are from the United States, including the National Institutes of Health and the US National Library of Medicine. The disparity in the number of studies in non-high-income countries is due to limited healthcare resources. This is significant considering that 80% of the global population resides in developing countries, where public health issues continue to rise due to rapid globalization and urbanization. Therefore, studies in developing countries are crucial, as the lack of data in these regions affects the applicability of findings to their specific contexts. Our study also identified that the most productive and prominent institutions funding AI research are from the United States, including the National Institutes of Health and the US National Library of Medicine. The disparity in the number of studies in non-high-income countries is due to limited healthcare resources. This is significant considering that 80% of the global population resides in developing countries, where public health issues continue to rise due to rapid globalization and urbanization. Therefore, studies in developing countries are crucial, as the lack of data in these regions affects the applicability of findings to their specific contexts.

### Predictors

Our study collected 250 different predictors from the 46 studies and grouped them into four categories: demographic (13 predictors; 5.2%), clinical (50 predictors; 20%), comorbidity (33 predictors; 13.2%), and laboratory (154 predictors; 61.6%). 42 (91.3%) studies included demographics, with the most used predictors being age, sex, and race. Demographics were included in 42 (91.3%) studies, with age, sex, and race being the most commonly used predictors. Clinical factors were included in 43 (93.5%) studies, featuring predictors like body mass index, blood pressure, and history of antidiabetic medication. Comorbidities were considered in only 23 (50%) studies, including hypertension, heart disease, and renal diseases. Laboratory parameters were utilized in 34 (73.9%) studies, with the most frequent predictors being HbA1c, high-density lipids, and cholesterol levels.

Testing an existing ML model in other settings needs to account for the availability of predictor data. In low-resource settings, a model which requires laboratory parameters might not be difficult to test due to limited infrastructure. We created a subgroup analysis of models with no laboratory data (i.e., demographic, clinical, or comorbidity), with only laboratory data, and mixed. The AUROCs of lab-only and non-lab models are 0.837 and 0.714 respectively. This means non-lab models were comparable and did not perform poorly compared to lab parameters.^22^ ^23^ To improve performance, several strategies can be employed, such as hyperparameter tuning and exploring different algorithms that can optimize the model.^24^

### Model Development

Most (n=29, 63.04%) included studies that utilized k-fold cross-validation as internal validation, similar to previous studies in diabetes risk prediction.^25^ With this method, the data is divided into k folds of equal size, and the model is subsequently trained and evaluated k times, with each evaluation utilizing a different fold as the test set. ^25^ This method is preferable compared to the hold-out approach as the whole dataset is utilized for development.^6^ ^25^ However, like other internal validation methods (including bootstrapping), optimism should be adjusted for in the final model.^6^

Only 21 (45.65%) studies reported how they handled missing data, although disregarding it may lead to imbalances, consequently introducing bias and misleading results. We found that models where missing data handling is described perform better. Reporting is essential as imputing different central tendencies (mean, median, or mode), as missing data could lead to different outcomes for different data distributions.^26^ More recently, autoencoders and other imputation techniques can more accurately fill in incomplete data.^27^ ^28^ These technologies would be beneficial for data pre-processing prior to AI model developments.

### Algorithm Types

Interestingly, our meta-analysis found gradient boosting to be the most common ML algorithm utilized, with a leading AUROC model performance of 0.789, followed by random forests (AUROC 0.776). Boosting algorithms are similar to random forests as they are ensemble learning algorithms, with the advantage of avoiding overfitting.^29^ ^30^ They also work well with categorical and numerical predictors. The third leading algorithm for performance, neural networks (AUROC 0.759), are comparatively less utilized by studies. As they fall in the deep learning category, despite their exceptional performance and capability to capture complex relationships, they are demanding computationally as they require large datasets.^30^

### External Validation

The uniform decrease of model performance when validated in external datasets (AUROC of 0.725) compared to internal validations (AUROC of 0.765) proved that development stages tend to overestimate, consistent with previous studies, such as non-AI prognostic model studies,^31^ or AI models for other purposes.^32^ ^33^ Moreover, only 11 (23.91%) of our included studies conducted external validation, despite it being a crucial step in prediction models and prognostic research, providing the capability for clinical impact over different settings.^34^ Contrarily, for some studies, such as those with small non-representative datasets or missing predictors, an external validation may not be worth it.^35^ Judging the overly optimistic nature of development models, diabetes complication prediction AI models may consider multiple external validations unless they are specifically made for local clinical settings.^36^

### Risk of Bias

An analysis of the risk of bias in published studies reveals significant issues in their design. First, many studies exhibit a high or unclear risk of bias, often due to incomplete data and insufficient population samplings, such as the underrepresentation of diverse patient groups and inadequate consideration of critical predictors like age and laboratory results—issues with data extraction, including incomplete or inconsistent datasets, further compromise model accuracy and reliability. Variable follow-up intervals also affect the generalizability of results.^37^

Second, The heavy reliance on internal validation with limited datasets from single centres is another concern, as studies lacking external validation show a higher risk of bias. In contrast, multi-center studies or those using national databases tend to have lower risk.^38^ The future success of machine learning prediction models hinges on high-quality, diverse training data. Proper data handling, capturing heterogeneity, and incorporating complexity are essential to enhance the models’ applicability and reliability.^39^

### Way Forward

The application of AI and machine learning (ML) in predicting complications is in its early stages, with significant potential due to the increasing complexity and volume of data. Early and accurate diagnosis of macrovascular complications could enable timely treatment, but this requires rigorous validation and scrutiny for effective outcomes. Enhancing reliability involves increasing external validation from diverse sources and promoting open-source development and interdisciplinary collaboration. While laboratory data enhances predictive accuracy, reliance on such data may limit the applicability of AI models in low-resource settings where lab facilities are not readily available. Future research should aim to improve the predictive power of non-lab models by incorporating advanced techniques for history-taking and physical examination data.

To ensure ethical and practical AI/ML use in healthcare, it is crucial to establish a secure framework focusing on data protection, secure handling, patient consent, and algorithmic transparency. Addressing biases and limitations is essential for broader implementation. Collaboration with policymakers, bioethicists, academics, and the broader community will be vital. Future research should prioritize validation and implementation strategies to improve the practical utility and trustworthiness of AI/ML models in clinical settings. Moreover, training healthcare professionals to interpret AI-driven predictions and incorporate them into patient management plans will further enhance the practical utility of these tools. Finally, continuous monitoring and updating of AI models with new data will ensure their ongoing accuracy and relevance in a rapidly evolving healthcare landscape.

### Strengths and Limitations

With the numerous models included in the meta-analysis, we are confident that this study reflects the capability of artificial intelligence in predicting diabetes complications to date. Information technology literature, such as IEEE Xplore, yielded studies from computer science fields that would have been absent in health-related databases. The included studies were done in multiple countries of varying income levels. Additionally, the detailed subgroup analysis provides valuable insights into the factors affecting model performance, such as the type of predictors (lab vs. non-lab) and the machine learning algorithms used.

Nevertheless, despite all relevant subgroup analyses explored, our meta-analysis has high heterogeneity, which can stem from differences in study populations, data sources, and machine learning algorithms used; such a phenomenon is commonly observed in published AI model performance meta-analyses.^33^ ^40^ Only a tiny proportion of the included studies conducted external validation, a crucial step for assessing the generalizability of prediction models. This lack of external validation raises concerns about the models’ applicability in different clinical settings. We also only analyzed AUROCs in our meta-analyses as the most utilized parameter; consequently, we excluded studies using different model performance parameters. Finally, the reliance on laboratory data for superior predictive accuracy may limit the practical implementation of these models in low-resource settings, where such data may not be readily available. Future research should focus on enhancing the performance of non-lab-based models to increase their applicability across diverse healthcare environments.

### Conclusions

This review demonstrates the promising potential of machine learning (ML) models in predicting macrovascular complications among individuals with type 2 diabetes mellitus (T2DM). We reveal a moderate overall performance with significant insights into the factors influencing predictive accuracy. However, the high heterogeneity observed among the included studies highlights the variability in model performance, emphasizing the need for tailored approaches based on specific populations and settings. Future studies should focus on developing robust non-lab-based models and conducting extensive external validations to improve the applicability of AI models in diverse clinical settings, especially in low-resource environments. Ultimately, the successful integration of AI and ML models in predicting diabetes complications will require interdisciplinary collaboration, ethical considerations, and ongoing validation to ensure their reliability and effectiveness in real-world clinical practice.

## Author Contributions

**Aqsha Nur**: Conceptualization, review protocol, article screening, data extraction, bias assessment, writing – original draft; **Sydney Tjandra**: Search strategy, article screening, data extraction, bias assessment, quantitative analysis and synthesis, writing – original draft; **Defin Yumnanisha**: data extraction, bias assessment, writing – original draft; **Arnold Keane**: data extraction, bias assessment, writing – original draft, writing – review and editing; **Adang Bachtiar**: Conceptualization, review protocol, writing – review.

## Author Contributions

Stevano Wijoyo and Rizqi Humaira (University of Indonesia) supported the initial screening of this review. Dante Harbuwono (University of Indonesia) provided feedback on the clinical impact of AI for diabetes, which guided the protocol of this review.

## Acknowledgments

Stevano Wijoyo (University of Indonesia) supported the initial screening of this review. Dante Harbuwono (University of Indonesia) and Sri Laksmiastuti (University of Trisakti) suggested the clinical impact of AI on diabetes, which guided the protocol of this review.

## Data Availability Statement

Data of this study are publicly available on the Open Science Framework, a public, open access repository, at https://osf.io/7gh9m/. Please contact the corresponding author for any further inquiries.

