## Supplementary Table for "Predicting the risks for stroke, cardiovascular disease, and peripheral vascular disease among people with type 2 diabetes with artificial intelligence models: a systematic review and meta-analysis"

### **Supplementary Material 1. PRISMA Checklist**

| **Section and topic** | **Item #** | **Checklist item** | **Reported on Page #** |
| --- | --- | --- | --- |
| **ADMINISTRATIVE INFORMATION** | | |  |
| **Title** |  |  |  |
| Title | [#1](https://www.goodreports.org/reporting-checklists/prisma/info/#1) | Identify the report as a systematic review | 1 |
| **Abstract** |  |  |  |
| Abstract | [#2](https://www.goodreports.org/reporting-checklists/prisma/info/#2) | Report an abstract addressing each item in the PRISMA 2020 for Abstracts checklist | 1 |
| **Introduction** |  |  |  |
| Background/rationale | [#3](https://www.goodreports.org/reporting-checklists/prisma/info/#3) | Describe the rationale for the review in the context of existing knowledge | 3 |
| Objectives | [#4](https://www.goodreports.org/reporting-checklists/prisma/info/#4) | Provide an explicit statement of the objective(s) or question(s) the review addresses | 3-4 |
| **Methods** |  |  |  |
| Eligibility criteria | [#5](https://www.goodreports.org/reporting-checklists/prisma/info/#5) | Specify the inclusion and exclusion criteria for the review and how studies were grouped for the syntheses | 4 |
| Information sources | [#6](https://www.goodreports.org/reporting-checklists/prisma/info/#6) | Specify all databases, registers, websites, organisations, reference lists, and other sources searched or consulted to identify studies. Specify the date when each source was last searched or consulted | 4 |
| Search strategy | [#7](https://www.goodreports.org/reporting-checklists/prisma/info/#7) | Present the full search strategies for all databases, registers, and websites, including any filters and limits used | 4 |
| Selection process | [#8](https://www.goodreports.org/reporting-checklists/prisma/info/#8) | Specify the methods used to decide whether a study met the inclusion criteria of the review, including how many reviewers screened each record and each report retrieved, whether they worked independently, and, if applicable, details of automation tools used in the process | 4 |
| Data collection process | [#9](https://www.goodreports.org/reporting-checklists/prisma/info/#9) | Specify the methods used to collect data from reports, including how many reviewers collected data from each report, whether they worked independently, any processes for obtaining or confirming data from study investigators, and, if applicable, details of automation tools used in the process | 4-5 |
| Data items | [#10a](https://www.goodreports.org/reporting-checklists/prisma/info/#10a) | List and define all outcomes for which data were sought. Specify whether all results that were compatible with each outcome domain in each study were sought (for example, for all measures, time points, analyses), and, if not, the methods used to decide which results to collect | 5 |
| Study risk of bias assessment | [#11](https://www.goodreports.org/reporting-checklists/prisma/info/#11) | Specify the methods used to assess risk of bias in the included studies, including details of the tool(s) used, how many reviewers assessed each study and whether they worked independently, and, if applicable, details of automation tools used in the process | 5 |
| Effect measures | [#12](https://www.goodreports.org/reporting-checklists/prisma/info/#12) | Specify for each outcome the effect measure(s) (such as risk ratio, mean difference) used in the synthesis or presentation of results | 5 |
| Synthesis methods | [#13a](https://www.goodreports.org/reporting-checklists/prisma/info/#13a) | Describe the processes used to decide which studies were eligible for each synthesis (such as tabulating the study intervention characteristics and comparing against the planned groups for each synthesis (item #5)) | 4 |
| Synthesis methods | [#13b](https://www.goodreports.org/reporting-checklists/prisma/info/#13b) | Describe any methods required to prepare the data for presentation or synthesis, such as handling of missing summary statistics or data conversions | 5 |
| Synthesis methods | [#13c](https://www.goodreports.org/reporting-checklists/prisma/info/#13c) | Describe any methods used to tabulate or visually display results of individual studies and syntheses | 5 |
| Synthesis methods | [#13d](https://www.goodreports.org/reporting-checklists/prisma/info/#13d) | Describe any methods used to synthesise results and provide a rationale for the choice(s). If meta-analysis was performed, describe the model(s), method(s) to identify the presence and extent of statistical heterogeneity, and software package(s) used | 5 |
| Synthesis methods | [#13e](https://www.goodreports.org/reporting-checklists/prisma/info/#13e) | Describe any methods used to explore possible causes of heterogeneity among study results (such as subgroup analysis, meta-regression) | 5 |
| Synthesis methods | [#13f](https://www.goodreports.org/reporting-checklists/prisma/info/#13f) | Describe any sensitivity analyses conducted to assess robustness of the synthesised results | 5 |
| Reporting bias assessment | [#14](https://www.goodreports.org/reporting-checklists/prisma/info/#14) | Describe any methods used to assess risk of bias due to missing results in a synthesis (arising from reporting biases) | 5 |
| Certainty assessment | [#15](https://www.goodreports.org/reporting-checklists/prisma/info/#15) | Describe any methods used to assess certainty (or confidence) in the body of evidence for an outcome | 5 |
| Data items | [#10b](https://www.goodreports.org/reporting-checklists/prisma/info/#10b) | List and define all other variables for which data were sought (such as participant and intervention characteristics, funding sources). Describe any assumptions made about any missing or unclear information | 5 |
| **Results** |  |  |  |
| Study selection | [#16a](https://www.goodreports.org/reporting-checklists/prisma/info/#16a) | Describe the results of the search and selection process, from the number of records identified in the search to the number of studies included in the review, ideally using a flow diagram (<http://www.prisma-statement.org/PRISMAStatement/FlowDiagram>) | 6 |
| Study selection | [#16b](https://www.goodreports.org/reporting-checklists/prisma/info/#16b) | Cite studies that might appear to meet the inclusion criteria, but which were excluded, and explain why they were excluded | 5-6 |
| Study characteristics | [#17](https://www.goodreports.org/reporting-checklists/prisma/info/#17) | Cite each included study and present its characteristics | 5-6 |
| Risk of bias in studies | [#18](https://www.goodreports.org/reporting-checklists/prisma/info/#18) | Present assessments of risk of bias for each included study | 7 |
| Results of individual studies | [#19](https://www.goodreports.org/reporting-checklists/prisma/info/#19) | For all outcomes, present for each study (a) summary statistics for each group (where appropriate) and (b) an effect estimate and its precision (such as confidence/credible interval), ideally using structured tables or plots | Supp Material 2 |
| Results of syntheses | [#20a](https://www.goodreports.org/reporting-checklists/prisma/info/#20a) | For each synthesis, briefly summarise the characteristics and risk of bias among contributing studies | 7-8 |
| Results of syntheses | [#20b](https://www.goodreports.org/reporting-checklists/prisma/info/#20b) | Present results of all statistical syntheses conducted. If meta-analysis was done, present for each the summary estimate and its precision (such as confidence/credible interval) and measures of statistical heterogeneity. If comparing groups, describe the direction of the effect | 7-8 |
| Results of syntheses | [#20c](https://www.goodreports.org/reporting-checklists/prisma/info/#20c) | Present results of all investigations of possible causes of heterogeneity among study results | 12 |
| Results of syntheses | [#20d](https://www.goodreports.org/reporting-checklists/prisma/info/#20d) | Present results of all sensitivity analyses conducted to assess the robustness of the synthesised results | 11 |
| Risk of reporting biases in syntheses | [#21](https://www.goodreports.org/reporting-checklists/prisma/info/#21) | Present assessments of risk of bias due to missing results (arising from reporting biases) for each synthesis assessed | N/A |
| Certainty of evidence | [#22](https://www.goodreports.org/reporting-checklists/prisma/info/#22) | Present assessments of certainty (or confidence) in the body of evidence for each outcome assessed | N/A |
| **Discussion** |  |  |  |
| Results in context | [#23a](https://www.goodreports.org/reporting-checklists/prisma/info/#23a) | Provide a general interpretation of the results in the context of other evidence | 12-15 |
| Limitations of included studies | [#23b](https://www.goodreports.org/reporting-checklists/prisma/info/#23b) | Discuss any limitations of the evidence included in the review | 16 |
| Limitations of the review methods | [#23c](https://www.goodreports.org/reporting-checklists/prisma/info/#23c) | Discuss any limitations of the review processes used | 16 |
| Implications | [#23d](https://www.goodreports.org/reporting-checklists/prisma/info/#23d) | Discuss implications of the results for practice, policy, and future research | 15 |
| **Other information** |  |  |  |
| Registration and protocol | [#24a](https://www.goodreports.org/reporting-checklists/prisma/info/#24a) | Provide registration information for the review, including register name and registration number, or state that the review was not registered | 4 |
| Registration and protocol | [#24b](https://www.goodreports.org/reporting-checklists/prisma/info/#24b) | Indicate where the review protocol can be accessed, or state that a protocol was not prepared | 4 |
| Registration and protocol | [#24c](https://www.goodreports.org/reporting-checklists/prisma/info/#24c) | Describe and explain any amendments to information provided at registration or in the protocol | N/A |
| Support | [#25](https://www.goodreports.org/reporting-checklists/prisma/info/#25) | Describe sources of financial or non-financial support for the review, and the role of the funders or sponsors in the review | 17 |
| Competing interests | [#26](https://www.goodreports.org/reporting-checklists/prisma/info/#26) | Declare any competing interests of review authors | 17 |
| Availability of data, code, and other materials | [#27](https://www.goodreports.org/reporting-checklists/prisma/info/#27) | Report which of the following are publicly available and where they can be found: template data collection forms; data extracted from included studies; data used for all analyses; analytic code; any other materials used in the review | 17 |

Supplementary Material 2. Search strategy keywords and hits for each database

Table 1: Search strategy keywords and hits for each database

| **Database** | **Search Strategy** | **Hits** |
| --- | --- | --- |
| **PubMed/ MEDLINE** | ("type 2 diabet*" OR T2D OR T2DM OR NIDDM OR "non-insulin dependent diabet*" OR "noninsulin-dependent diabet*" OR "maturity-onset diabet*" OR "maturity onset diabet*" OR "adult-onset diabet*" OR "adult onset diabet*" OR MODY OR "noninsulin dependent diabet*" OR "non-insulin-dependent diabet*") AND ("artificial intelligence" OR "machine learning" OR AI OR "knowledge bas*" OR "neural net*" OR "network model" OR "deep learning" OR "data mining" OR "fuzzy logic" OR "expert systems" OR "decision tree" OR "random forest" OR "nearest neighb*" OR "support vector machin*" OR "gbm" OR "gradient boosting") AND ("prediction*" OR "predictive modelling" OR "predictive models" OR "predictor*" OR "score") AND ("risk" OR "event" OR "inciden*" OR "complication*") AND (stroke[MeSH] OR cardiovascular disease[MeSH] OR "CVD" OR "ischemic heart disease" OR "coronary artery" OR "heart failure" OR "peripheral arterial disease*" OR "peripheral artery disease*" OR peripheral vascular disease[MeSH] OR "limb ischemia" OR "limb ischaemia" OR "PAD") | 245 |
| **Scopus** | ("type 2 diabet*" OR T2D OR T2DM OR NIDDM OR "non-insulin dependent diabet*" OR "noninsulin-dependent diabet*" OR "maturity-onset diabet*" OR "maturity onset diabet*" OR "adult-onset diabet*" OR "adult onset diabet*" OR MODY OR "noninsulin dependent diabet*" OR "non-insulin-dependent diabet*") AND ("artificial intelligence" OR "machine learning" OR AI OR "knowledge bas*" OR "neural net*" OR "network model" OR "deep learning" OR "data mining" OR "fuzzy logic" OR "expert systems" OR "decision tree" OR "random forest" OR "nearest neighb*" OR "support vector machin*" OR "gbm" OR "gradient boosting") AND ("prediction*" OR "predictive modelling" OR "predictive models" OR "predictor*" OR "score") AND ("risk" OR "event" OR "inciden*" OR "complication*") AND ("stroke" OR "cardiovascular disease" OR "CVD" OR "ischemic heart disease" OR "coronary artery" OR "heart failure" OR "peripheral arterial disease*" OR "peripheral artery disease*" OR "peripheral vascular disease*" OR "limb ischemia" OR "limb ischaemia" OR "PAD") | 344 |
| **Google Scholar** | diabetes mellitus and artificial intelligence and complication and stroke \| cardiovascular \| peripheral disease | 100 |
| **IEEE Xplore** | ("All Metadata":diabetes) AND ("All Metadata":artificial intelligence OR "All Metadata":machine learning OR "All Metadata":AI OR "All Metadata":knowledge bas* OR "All Metadata":neural net* OR "All Metadata":network model OR "All Metadata":deep learning OR "All Metadata":data mining OR "All Metadata":fuzzy logic OR "All Metadata":expert systems OR "All Metadata":decision tree OR "All Metadata":random forest OR "All Metadata":nearest neighb* OR "All Metadata":support vector machin* OR "All Metadata":gbm OR "All Metadata":gradient boosting) AND ("All Metadata":stroke OR "All Metadata":cardiovascular OR "All Metadata":coronary artery OR "All Metadata":heart OR "All Metadata":peripheral arterial OR "All Metadata":peripheral vascular) AND ("All Metadata":prediction OR "All Metadata":predictive modelling OR "All Metadata":predictive model* OR "All Metadata":predictor* OR "All Metadata":score) AND ("All Metadata":risk OR "All Metadata":event OR "All Metadata":inciden* OR "All Metadata":complication*) | 219 |
| **EMBASE** | ('type 2 diabet*' OR t2d OR 't2dm'/exp OR t2dm OR 'niddm'/exp OR niddm OR 'non-insulin dependent diabet*' OR 'noninsulin-dependent diabet*' OR 'maturity-onset diabet*' OR 'maturity onset diabet*' OR 'adult-onset diabet*' OR 'adult onset diabet*' OR mody OR 'noninsulin dependent diabet*' OR 'non-insulin-dependent diabet*') AND ('artificial intelligence'/exp OR 'artificial intelligence' OR 'machine learning'/exp OR 'machine learning' OR ai OR 'knowledge bas*' OR 'neural net*' OR 'network model'/exp OR 'network model' OR 'deep learning'/exp OR 'deep learning' OR 'data mining'/exp OR 'data mining' OR 'fuzzy logic'/exp OR 'fuzzy logic' OR 'expert systems'/exp OR 'expert systems' OR 'decision tree'/exp OR 'decision tree' OR 'random forest'/exp OR 'random forest' OR 'nearest neighb*' OR 'support vector machin*' OR 'gbm' OR 'gradient boosting'/exp OR 'gradient boosting') AND ('prediction*' OR 'predictive modelling'/exp OR 'predictive modelling' OR 'predictive models' OR 'predictor*' OR 'score'/exp OR 'score') AND ('risk'/exp OR 'risk' OR 'event' OR 'inciden*' OR 'complication*') AND ('stroke'/exp OR 'stroke' OR 'cardiovascular disease'/exp OR 'cardiovascular disease' OR 'cvd' OR 'ischemic heart disease'/exp OR 'ischemic heart disease' OR 'coronary artery'/exp OR 'coronary artery' OR 'heart failure'/exp OR 'heart failure' OR 'peripheral arterial disease*' OR 'peripheral artery disease*' OR 'peripheral vascular disease*' OR 'limb ischemia' OR 'limb ischaemia' OR PAD) | 688 |
| **Wiley Onlline Library** | ("type 2 diabet*" OR T2D OR T2DM OR NIDDM OR "non-insulin dependent diabet*" OR "noninsulin-dependent diabet*" OR "maturity-onset diabet*" OR "maturity onset diabet*" OR "adult-onset diabet*" OR "adult onset diabet*" OR MODY OR "noninsulin dependent diabet*" OR "non-insulin-dependent diabet*") AND ("artificial intelligence" OR "machine learning" OR AI OR "knowledge bas*" OR "neural net*" OR "network model" OR "deep learning" OR "data mining" OR "fuzzy logic" OR "expert systems" OR "decision tree" OR "random forest" OR "nearest neighb*" OR "support vector machin*" OR "gbm" OR "gradient boosting") AND ("prediction*" OR "predictive modelling" OR "predictive models" OR "predictor*" OR "score") AND ("risk" OR "event" OR "inciden*" OR "complication*") AND ("stroke" OR "cardiovascular disease" OR "CVD" OR "ischemic heart disease" OR "coronary artery" OR "heart failure" OR "peripheral arterial disease*" OR "peripheral artery disease*" OR "peripheral vascular disease*" OR "limb ischemia" OR "limb ischaemia" OR PAD) | 1421 |

### Supplementary Material 3. PICOT Inclusion and Exclusion Criteria

Table 2: PICOT inclusion and exclusion criteria

|  | **Participants (P)** | **Intervention (I)** | **Comparison (C)** | **Outcomes (O)** | **Timeframe (T)** | **Others** |
| --- | --- | --- | --- | --- | --- | --- |
| **Inclusion Criteria** | Adults with type 2 diabetes mellitus; actual medical records/datasets | AI development and implementation, including machine learning and deep learning | N/A | Prediction performances for stroke, cardiovascular disease, or peripheral vascular disease | Between January 1, 2000, to November 30, 2023 | Diagnostic or prognostic studies; English language |
| **Exclusion Criteria** | Mixed populations with other types of diabetes or prediabetes; non-human subjects | Classical statistical models, e.g. logistic regression, without specific mention of AI; drug involvement |  | Prediction performances for diabetes; qualitative studies |  | Irretrievable full-text; theoretical models; reviews; framework developments; proposals |

### Supplementary Material 4. General study characteristics

Table 3: General study characteristics

| No | Author (Year) | Country (Income) | Data sources | N | Predictors | Algorithm | Outcome | Handling Missing Data | Cross-validation (...fold) | External Validation |
| --- | --- | --- | --- | --- | --- | --- | --- | --- | --- | --- |
| 1 | Nanda (2022) | India (LMIC) | Recruited patients from outpatient department | 160 | **Demographic** (Gender, age)  **Clinical** (weight, BMI, duration of T2DM, on insulin, smokers, blood pressure)  **Comorbidities** (hypertension, neuropathy, retinopathy)  **Laboratory** (fasting plasma glucose, postprandial plasma glucose (PPPG), HbA1c, urea, creatinine, uric acid, cholesterol,triglycerides, HDL, LDL, ApoA1, vitamin B12, TNF-alpha, IL-10) | Random forest, Neural network (KNN), Gradient boosting (AdaBoost), Others (SVM-PolyK, SVM-RBF, Naive-Bayes, Bagging, Stacking (RF + KNN)) | Diabetic foot ulcer type | N/A | 10 | No |
| 2 | Senthilkumar, 2023 | India (LMIC) | Cleveland & diabetes dataset; UCI repository (available online) | 303 | **Demographic** (age, sex)  **Clinical** ( chest pain type, resting blood pressure, resting ECG, maximum heart rate,exercise-induced angina, ST depression induced by exercise, slope of peak exercise ST segment, Significant vessels coloured by fluoroscopy, defect, angiographic disease status)  **Laboratory** (serum cholesterol, fasting blood sugar) | Logistic regression, Random Forest, Gradient Boosting (AdaBoost), Others (Multilayer perceptron). | CVD | Imputation | 10 | No |
| 3 | Sonia, 2023 | India (LMIC) | PhysioNet database, Southeastern Americans | 10344 | **Clinical** (Heart rate variability features: PR, LQT, AF, AFL, LBBB, QAb, TAb, LPR, VPB, LQRSV, IAVB, PAC, LAD, SB, Brady, SNR, STach, PVC, SA, LAnFB, RAD, RBBB, TInv, SVPB, NSIVCB, IRBBB, CRBBB) | Neural networks (DNN, AlexNet, LeNet-5, DNHRV) Others (Inception, VGG-16, LSTM) | CVD | Not applicable | Yes; N/A | No |
| 4 | Ding, 2023 | China (UMIC) | 3BExt database (hospital EHR) | 4722 | **Demographic** (age, sex, classification of chinese hospitals, education level, race, state of motion, geographical regions, health insurance, marital status)  **Clinical** (BMI, waist, height, weight, diabetes duration, smoking for 5 years, no smoking for 5 years, compliance, blood pressure, antidiabetic drug, antihypertensive drug, hypolipidemic agents, family history of diabetes)  **Comorbidities (**Cardiovascular disease; history of other cardiovascular diseases, diabetic peripheral vascular disease, history of microvascular complications, hypoglycemia)  **Laboratory** (Total cholesterol, TG, HbA1c, LDL, FPG, HDL) | Logistic regression, Gradient Boosting [AdaBoost, Gradient boosting decision tree (GBDT)], Random Forest, Others (SVM) | ASCVD (CAD,CeVD, PVD) | Imputation | N/A | Yes |
| 5 | Abegaz, 2023 | United States (HIC) | AoU research data v6 | 9059 | **Demographic (**race, gender)  **Clinical** (DBP, SBP, BMI)  **Laboratory** (Phosphate, chloride, potassium, Sodium, triglyceride, creatinine, TCHOL, BUN, HBA1C, neutrophils, monocytes, basophils, HDL, urate, ACR, hemoglobin, calcium, CRP, GFR, MCV, LDH, BNP, MCHC, Myoglobin, WC) | Random Forest, Gradient Boosting (XGBoost), Logistic Regression, Others (Weighted ensemble model) | MACE, MI, HF, Stroke | Imputation if <25% missing; Excluded if >25% | Yes; N/A | No |
| 6 | Selvarathi, 2023 | India (LMIC) | HippoKraion general hospital medical records | 560 | **Demographic** (age, sex)  **Clinical** (BMI, smoking habit, fibrates, statins, aspirin, insulin usage, pulse pressure, diabetes duration, parental history of DM)  **Comorbidities (**hypertension)  **Laboratory**  (fasting glucose, HDL cholesterol, triglycerides, HbA1c) | Neural network (Ensemble hybrid wavelet neural network (HWNN)), Others (CCGLSTM,Traditional LSTM, Convolution LSTM, Convolution GLSTM) | CVD | Imputation | Yes | No |
| 7 | Vimont, 2023 | France (HIC) | French National Health Data Information System | 22708 | **Demographic** (Age)  **Clinical** (disease duration, diabetes medications)  **Comorbidities (**chronic cardiovascular disease, psychiatric disorder, cancer, chronic end-stage renal disease, neurologic disorder, COPD, inflammatory or rare disease, liver or pancreas disease) | Logistic Regression, Random Forest, Neural network | CVD (HF, PAD, MI, Stroke, UA, TIA, CV-related death) | N/A | 100 | Yes |
| 8 | Gandin, 2023 | Italy (HIC) | Cardiovascular Observatory of Trieste (Italy) EHR | 10614 | **Demographic** (Age)  **Clinical** (BMI, LV WMSI, P axis absent, Aortic max vel CW, Diuretics (other), T axis, Anticoagulant, LV E' vel, P axis)  **Laboratory** (Hemoglobin, Glycemia, Triglyceride)  **Comorbidities** (Renal disease, Hypertension, Lung disease, Tricuspid regurgitation, RASi, Pericardium disease, Peripheral arterial disease) | Cox-based (Cox proportional hazards regression), Neural Network (Non-linear proportional hazards deep neural network (PHNN)) | HF | Imputation | 10 | No |
| 9 | Zhong, 2022 | China (UMIC) | Shaoxing People’s Hospital EHR | 521 | **Demographic** (Sex, age)  **Clinical** (history of smoking, drinking, espiratory rate, heart rate, SBP, DBP, Killip grade, family history of CVD)  **Comorbidities** (hypertension, hyperlipidemia)  **Laboratory** (Serum biomarkers including aspartate aminotransferase (AST), lactate dehydrogenase, total bilirubin, total protein, albumin, globulin, albumin/globulin ratio, urea, creatinine, uric acid, total cholesterol, triglyceride (TG), high-density lipoprotein, low-density lipoprotein cholesterol, apolipoprotein A1, apolipoprotein B, apolipoprotein B/apolipoprotein A1, fasting blood glucose (FBG), α-hydroxybutyrate dehydrogenase, creatine kinase MB, homocysteine, C-reactive protein, neutrophil count, lymphocyte count, neutrophil-lymphocyte ratio, HbA1c, and triglyceride-glucose (TyG) index) | LR, LR with lasso, KNN, SVM linear, SVM radial, Decision tree, Random forest, Gradient Boosting (XGBoost), Neural Network (ANN), Logistic Regression (LR with Lasso), Others (SVM linear, SVM radial, Decision tree) | ACS | Not applicable | 5 | Yes |
| 10 | Miran, 2021 | United States (HIC) | Health Facts database | 723 | **Demographic** (Age) **Comorbidities** (hypertension, ischemic heart disease, atrial fibrillation, chronic kidney disease, anemia, arthritis, asthma, COPD, cancer, depression) | Logistic Regression, Neural network, Random Forest, Gradient Boosting (XGBoost) | HF | N/A | 5 | No |
| 11 | Momenzadeh, 2022 | United States (HIC) | Clinical research data cohort warehouse | 4646 | **Demographic** (sex, marital status, employment status, race, age at diabetes diagnosis)  **Clinical** (BMI, DBP, SBP, pulse, temperature, respiration rate, alcohol use, illicit drug use, tobacco use)  **Laboratory** (aspartate aminotransferase, alanine transaminase, bilirubin, alkaline phosphatase, calcium, glucose, bicarbonate, chloride, sodium, potassium, creatinine, eGFR, BUN, anion gap, platelet count, hematocrit, Hb, RBC count, white blood cell count, MCHC, MCV, MPV, RDW, monocyte percentage, neutrophil percentage, eosinophil percentage, lymphocyte percentage, absolute neutrophil count, absolute lymphocyte count, absolute monocyte count, absolute eosinophil count, total protein, albumin) | Gradient Boosting (AdaBoost, XGBoost) Others (SVC, ET), Random Forest, Logistic Regression | CVD | MissForest; excluded if 50% or more values were missing | 5 | No |
| 12 | Nicolucci, 2022 | Italy (HIC) | EMR | 147664 | **Demographic** (gender, year of birth, date of diagnosis)  **Clinical** (BMI, height, weight, waist circumference, SBP, DBP, ABI DX, ABI SX)  **Laboratory** (Albuminuria, serum creatinine, Creatinine clearance, Total cholesterol, LDL HDL, Triglycerides, Fibrinogen, GGT, ALT AST, Alkaline phosphatase Amylase CPK Hemoglobin Platelets BUN Uric acid Glycosuria Urinary amylase Urinary ketones Urinary potassium Urinary sodium Urine creatinine Urine culture (positive/negative)) | Gradient Boosting (XGBoost) | CVD | Imputation | 10 | Yes |
| 13 | Hong, 2021 | United States (HIC) | EHR | 6245 | **Demographic** (age at diabetes diagnosis, race/ethnicity, sex)  **Clinical** (BMI, weight, height, blood pressure, diagnoses of various diseases, drugs)  **Laboratory** (total cholesterol, triglycerides, high-density lipoprotein (HDL) cholesterol, low-density lipoprotein (LDL) cholesterol, glycosylated hemoglobin (HbA1c) and estimated glomerular filtration rate (eGFR)) | Cox-based (Cox proportional hazard), Others (LASSO regression) | CHD, HF, Stroke | Excluded | N/A | No |
| 14 | Aminian, 2020 | United States (HIC) | EHR | 2287 | **Demographic** (sex, age, race, zip code income, location)  **Clinical** (BMI, smoking status, medical history of diabetes medication / insulin / lipid-lowering / RAA system inhibitors / other antihypertensive / aspirin / warfarin, SBP, DBP)  **Laboratory** (HbA1c, eGFR, HDL, LDL, Triglycerides, UACR) | Cox-based (Cox proportional hazards, exponential, Fine-Gray), Random Forest | Coronary artery events | Multivariate imputation by chained equations | 5 | No |
| 15 | Athanasiou, 2020 | Greece (HIC) | Hospital EHR | 560 | **Demographic** (Age)  **Clinical** (BMI, smoking, parental history, diabetes duration, blood pressure, lipid lowering therapy)  **Comorbidity** (Hypertension)  **Laboratory** (HDL, HbA1c, triglycerides, total cholesterol, fasting glucose) | Gradient Boosting (XGBoost) | CVD | N/A | 10 | No |
| 16 | Miao, 2020 | China (UMIC) | Framingham Heart Study dataset | 8391 | **Demographic** (age)  **Clinical (**BMI)  **Laboratory** (fasting plasma glucose) | Neural network (KNN), Others (SVM) | CVD | N/A | N/A | No |
| 17 | Hossain, 2021 | Australia (HIC) | CBHS Health Fund Australia | 172 | **Demographic** (age, sex)  **Comorbidity (**disease network (comorbidities)) | Logistic Regression, Others (SVM, Decision Tree, NB), Random Forest, Neural Network (KNN). | CVD | N/A | 10 | No |
| 18 | Zarkogianni, 2018 | Greece (HIC) | Hospital medical records | 560 | **Demographic** (Age, Sex)  **Clinical (**Duration of Diabetes, Smoking Habit, HbA1c, Systolic Blood Pressure)  **Laboratory** (Total cholesterol, HDL cholesterol) | Random Forest, Others (Hybrid ensemble, SOM classifier, BLR model, CART, NB), Neural network (FFN) | CHD, Stroke | N/A | 10 | No |
| 19 | Segar, 2019 | United States (HIC) | ACCORD Trial Dataset | 8756 | **Demographic** (Age)  **Clinical** (BMI, QRS duration, MI, and CABG)  **Comorbodity** (Hypertension)  **Laboratory** (Creatinine, HDL-C, fasting plasma glucose) | Random survival forest (RSF), Cox-based method | HF | Random forest imputation | N/A | Yes |
| 20 | Derevitskii, 2020 | Russia (UMIC) | V.A. Almazov National Medical Research Center | 8139 | **Demographic** (Age)  **Clinical** (BMI, diabetic retinopathy, insulinum glarginum, arterial hypertension, weight, angina pectoris, atrial fibrilation, atherosclerosis, coronary heart disease) | Gradient Boosting (XGBoost) | CHF, AF | N/A | N/A | No |
| 21 | Ljubic, 2020 | United States (HIC) | the HCUP State Inpatient Databases of California database. | 1910674 | **Clinical** (Number of hospitalizations) | Neural Network (Recurrent neural network (RNN) RNN gated recurrent unit (GRU)) | Angina pectoris, Atherosclerosis, ICHD (ischemic chronic heart disease), Myocardial infarction, PVD | N/A | Yes;N/A | No |
| 22 | Fan, 2020 | China (UMIC) | Recruited from Inpatient and Outpatient LuHe Hospital | 1273 | **Demographic** (Age)  **Clinical** (Course of diabetes, heart rate, diastolic pressure, course of hypertension)  **Laboratory (**LDL cholesterol, total cholesterol, blood platelet) | Random forest (RF) | CHD | N/A | 5 | Yes |
| 23 | Dalakleidi, 2013 | Greece (HIC) | Medical Record of General Hospital Athens | 560 | **Demographic** (Age, Gender)  **Clinical (**DBP, MAP, incidence of DM in parents, use of calcium antagonists, b-blockers, proteinuria, diguanides and insulin treatment)  **Laboratory** (HbA1c, blood glucose, cholesterol, HDL, LDL cholesterol, non HDL cholesterol) | Others (Genetic algorithm (GA)) | fatal and nonfatal CVD | N/A | 10 | No |
| 24 | Xu, 2017 | China (UMIC) | EHR | 111761 | **Demographic** (Age) **Laboratory (**Cholinesterase, MCHC, Lipoprotein(a)  Na, Bacteria, Lactate dehydrogenase, Prealbumin, AST Total protein, Uric acid) | Logistic Regression (LR) | Stroke | k-NN imputation | N/A | No |
| 25 | Lee, 2023 | South Korea (HIC) | Seoul St. Mary's Hospital EMR | 5040 | **Demographic** (Age, sex)  **Clinical** (Medication history, height, weight, blood pressure)  **Laboratory** (HbA1c, blood urea nitrogen (BUN), creatinine, AST, ALT, total cholesterol, triglycerides, HDL-C, and LDL-C) | Others (GRU-ODE-Bayes-based) | CVD | Imputation | 10 | No |
| 26 | Wang, 2023 | China (UMIC) | Enrolled patients, Department of Endocrinology, University of Chinese Academy of Science Shenzhen Hospital | 111 | **Laboratory** (Metabolites) | Random Forest (RF) | Diabetic foot | Not applicable | N/A | No |
| 27 | Ozturk, 2023 | United Kingdom (HIC) | Connected Brafrod Dataset (Survey) | 43000 | **Clinical** (BMI)  **Laboratory** (Neutrophil, serum cholesterol, platelet count, serum alkaline phosphatase, serum alanine aminotransferase, serum creatinine, total white blood count, mean cell volume, serum sodium, HbA1c, monocyte count, red blood cell, GFR, serum total bilirubin, serum urea, serum albumin, serum potassium, lymphocyte, eosinophil) | Neural networks (NN), Random Forest (RF), Others (NB, SVM, Ensemble (Generalized LInear Model)) | Hypertension | Imputation | N/A | No |
| 28 | Kanda, 2022 | Japan (HIC) | the Japan Medical Data Vision (MDV) databses | 217054 | **Demographic** (Age, sex)  **Clinical** (BMI, frequency of outpatient visit, and frequency of hospitalization)  **Laboratory** (lab tests) | Gradient Boosting (XGBoost), Neural Network, Logistic regression, Cox-based (Cox proportional hazard) | CVD | Imputation | N/A | Yes |
| 29 | Lee, 2021 | Hong Kong (HIC) | Clinical Data and Reporting System | 261308 | **Demographic** (Age, sex)  **Clinical (**Medication)  **Laboratory** (Liver function test, CBC, lipid profile, renal function test, glycemic control (HbA1c)) | Random Forest (Random survival forest), Cox-based (Multivariate Cox model), Others (Conditional inference survival forest (CISF)) | AMI | N/A | 5 | No |
| 30 | Liu, 2020 | United States (HIC) | Truven MarketScan Commercial Claims and Encounter (CCAE) database | 53275 | **Demographic** (Gender, Age)  **Clinical** **(**ICD-10 diagnoses, medications (19 classes related to glucose control, cardiac related drugs, and antibiotics) | Multi task learning (Multi-task feature learning (MTFL), Outcome-specific multi-task learning (OS-MTL), Private-shared MTL), Others (Single-task learning with Lasso) | Vascular Disease (peripheral, cardiovascular, or cerebral) | N/A | 10 | No |
| 31 | Farzi, 2017 | Iran (LMIC) | Hospital Medical Records | 1000 | **Demographic** (Gender, Age)  **Clinical** (Infection year, Blood pressure, Overweight, Smoking)  **Laboratory** (FBS, HBA1C, BS2HP, CR, CHO, TG, HDL, LDL, GLUCOSE, PROTEIN, WBC, RBC, Blood fat) | Random Forest, Others (J48 (Decision Tree), LMT, NBTree, SMO, MLP (Multi-Layer Perception neural network), Naïve Bayes, Bayes Net, and RBF (Radial Base Function). | CVD |  | N/A | No |
| 32 | Longato, 2020 | Italy (HIC) | Administrative Repository | 97466 | **Demographic** (Age,  gender)  **Clinical (**Estimated diabetes duration**)** | Neural network (NN) | CVD, Stroke | N/A | N/A | No |
| 33 | Giardina, 2006 | United Kingdom (HIC) | Ulster Hospital EHR | 352 | **Demographic** (Gender, BMI, Weight)  **Clinical** (Age at diagnosis, smoking status, systolic blood pressure, diastolic blood pressure)  **Laboratory** (total HDL-cholesterol ratio, low HDL-cholesterol,high LDL cholesterol,total cholesterol, HbA1c, urinary albumin/creatinine ratio, micro-albumin, creatinine, Random blood glucose, Triglycerides, Urea) | Neural network (Weighted k-nearest neighbors (WkNN), kNN), Others (Genetic algorithm (GA), Random initialization (RI)) | CHD | kNN imputation | N/A | No |
| 34 | Phan, 2023 | Taiwan (HIC) | Taipei Medical University Clinical Research Database | 39646 | **Demographic** **(**Age, gender)  **Clinical** (disease histories, medication use)  **Laboratory** (glucose, HbA1c, etc) | Logistic Regression (LR), Gradient Boosting (Gradient boosting machine (GBM), Light Gradient Boosting Machine (LGBM), AdaBoost, and eXtreme Gradient Boosting (XGB)), Random Forest (RF), Others (Voting ensemble, Linear discriminant analysis (LDA)) | Ischemic Stroke | Median Imputation | 5 | Yes |
| 35 | Rahman, 2018 | Bangladesh (LMIC) | Tokyo Women‘s Medical University Hospital | 779 | **Demographic** (Age, Sex)  **Clinical** (Height, Weight, Waist circumstances, Duration of T2DM, Blood Pressure (Systolic and Diastolic), Pulse rate, Weight, BMI, Insulin, Smoking habit, Drinking habit, Risk of Cardiovascular disease and Risk of Nephropathy)  **Comorbidity** (History of complications, History of Hypertension, History of Arteriosclerosis Obliterans, History of Atrial Fibrillation, History of Myocardial Infarction, History of Dyslipidemia, History of Myocardial Infarction, History of Cerebral Infarction, History of Angina Pectoris, History of Heart Failure, History of Retinopathy, History of Kidney disease, History of Hyperuricemia, History of Liver disease)  **Laboratory** (HbA1c, Fasting Plasma Glucose, HOMA-R, HOMA-beta, C-peptide, CPI, 1,5-anhydroglucitol,1,4 -anhydro (-D)-glucitol, Glycoalbumin, Proinsulin, Na, K, Cl, Triglyceride, HDL-C, LDL-C, BUN, Uric acid, Creatinine, eGFR, Red Blood, White Blood, Hemoglobin, Hematocrit, Platelets, AST, ALT, and γ-GTP, Urinary Protein and Urinary Sugar) | Logistic Regression (LR), Random Forest, Gradient Boosting (Decision Tree with AdaBoost), Others (SVM, NB, Decision Tree) | CVD | Imputation | 10 | No |
| 36 | Liu, 2018 | United States (HIC) | MarketScan Commercial Claims and Encounter (CCAE) database from Truven Health. | 8240 | **Demographic** (age, gender)  **Clinical** (weight index, ICD codes) | Multi-task learning, Others (Single-task learning (STL)) | Vascular Disease | N/A | N/A | No |
| 37 | Rajathi, 2020 | India (LMIC) | N/A | 770 | **Demographic** (Sex, age)  **Clinical**  (Smoking habit, chest pain, diabetes duration,BMI, pulse pressure)  **Laboratory** (glycosylated hemoglobin, fasting glucose) | Neural Network (Hybrid Wavelet Neural Network), Others (Self Organized Mapping, Modified Teaching Learning Based Optimization) | CVD | N/A | N/A | No |
| 38 | Liu, 2020 | China (UMIC) | National Health Clinical Center | 1485 | **Demographic** (Age, gender)  **Laboratory** (Urine leucocyte, specific gravity, urobilinogen, urine bilirubin, RBC, yeast-like cells, glucosuria, crystaluria, urine pH value, urine color, griess test, urine turbidity, urine ketone, HbA1c, ALT, AST, TP, ALB, TB, DBIL, ALP, urea, GGT, Creatinine, Triglycerides, Uric Acid, Total Cholesterol, Creatine Kinase, LDH, Ca, Na, K, Chloride, Inorganic Phosphorus, Mg, HDL) | Random Forest (RF), Others (Bayesian Network (BN), NB, C5.0) | Macrovascular Complications, Diabetic foot | Multiple Imputations | 10 | No |
| 39 | Dworzynski, 2020 | Denmark (HIC) | Danish National Patient Register | 203517 | **Demographic** (Age, sex, address informatio) **Clinical** (Date of diagnosis prescriptions, hospital diagnoses, procedures, primary care interactions) | Logistic Regression (Reference logistic regression, Logistic ridge regression), Random forest, Gradient boosting | HF, MI, stroke, CVD | N/A | 3 | Yes |
| 40 | Afarideh, 2016 | Iran (LMIC) | Diabetes Clinic Database, Health Surveillance Centers | 2244 | **Demographic** (Age, sex)  **Clinical** (BMI, waist circumference, lipid-lowering and antihyperglycemic medication use, family history of CVD, smoking status, hypertension  **Laboratory** (A1C, ALT, AST, aP, GGT, LDL-C,HDL-C, triglycerides levels) | Cox-based (Cox proportional hazards), Neural network (Artificial neural network (ANN)) | Cardiovascular Disease | N/A | N/A | No |
| 41 | Liu, 2018 | United States (HIC) | Market Scan Commercial Claims and Encounter (CCAE) database from Truven Health | N/A | **Demographic** (Age, gender)  **Clinical** (Diagnoses (Chronic ulcer of skin; Disorders of fluid, electrolyte, and acid-base balance; Hereditary and idiopathic peripheral neuropathy; Atherosclerosis), medications | Logistic regression, Multi-task learning (Single task learning (STL), Multi-task feature learning (MTFL), Multi-task relationship learning (MTRL), Feature and task relationship learning (FETR), Others (TREFLES) | Vascular Disease | N/A | 5 | No |
| 42 | Mei, 2019 | China (UMIC) |  | 54482 | **Demographic** (Gender, age, race)  **Clinical** (systolic blood pressure (SBP), antihypertensive treatment, diabetes, and smoking)  **Laboratory** (Total cholesterol (TC), high-density lipoproteins cholesterol (HDL-C)) | Logistic regression, Neural network (Teacher-student network (TSNN), Knowledge-enhanced neural network (KENN), Others (Decision fusion, Pooled cohort equations) | CVD | Excluded | N/A | No |
| 43 | Sierra-Sosa, 2019 | Spain (HIC) |  | 149015 | **Demographic** (Age, gender)  Comorbidities | Logistic Regression (LR Ridge, LR Lasso), Others (Linear discriminant analysis (LDA), SVM) | MI | N/A | 10 | No |
| 44 | Thomas, 2018 | United States (HIC) |  | 805867 | **Demographic** (Age, gender, race)  **Clinical** (metabolic measurements, diagnosis codes, medical procedures, prescriptions)  **Laboratory** (results from clinical laboratory tests) | Others (Patient network) | Stroke, MI, HF | Imputation by Pyspark | N/A | No |
| 45 | Kim 2019 | United States (HIC) |  | 970069 | **Demographic** (age, gender, census region)  **Clinical** (smoking status, vital signs (BP, pulse, and Body Mass Index BMI), use of drugs related to comorbid)  **Comorbidity** (preexisting complications (chronic kidney disease (CKD), chronic renal failure (CRF), ischemic heart disease (IHD), congestive heart failure (CHF), peripheral vascular disease (PVD), and cerebrovascular disease (CVD))  **Laboratory** (lab results (HbA1c, FPG, lipid panel, Glomerular Filtration Rate GFR) | Multi-task learning, Gradient Boosting (GBM), Others (LASSO) | IHD, CHF, CVD, PVD | Excluded | N/A | Yes |
| 46 | Kim 2018 | United States (HIC) |  | 9793 | **Demographic** (age, gender)  **Clinical** (smoking status, vital signs (BP, pulse, and Body Mass Index BMI))  **Comorbidity** (dyslipidemia, hypertension, obesity)  **Laboratory** (HbA1c, lipid panel, Glomerular Filtration Rate GFR)) | Multi-task learning, Cox-based (Lasso-penalized Cox-regression) | IHD, PVD, CHF, CVD | N/A | N/A | Yes |

#### outcome, validation, algorithm, country/country income, missingness, cross-validation, predictors

### Supplementary Material 5. Performance outcomes of included studies

Table 3: Performance outcomes of included studies

| Author (year) | Algorithm | Outcome | F measure | AUC [SE] / AUC [Range] | SE | Sensitivity (Recall) | Specitivity | Accuracy | Precision (PPV) |
| --- | --- | --- | --- | --- | --- | --- | --- | --- | --- |
| Nanda, 2022 | RF | Diabetic foot ulcer type | 0.7 | 0.918 [0.0229 |  | 0.95 | 0.875 | 0.705 | N/A |
|  | SVM-PolyK |  | 0.635 | 0.818 [ 0.0336 |  | 0.938 | 0.938 | 0.638 | N/A |
|  | SVM-RBF |  | 0.584 | 0.828 [0.0328 |  | 0.925 | 0.9 | 0.519 | N/A |
|  | KNN |  | 0.7 | 0.828 [0.0328 |  | 0.825 | 0.85 | 0.718 | N/A |
|  | Naive-Bayes |  | 0.55 | 0.836 [0.0321] |  | 0.875 | 0.925 | 0.564 | N/A |
|  | AdaBoost |  | 0.268 | 0.662 [0.0428] |  | 0.813 | 0.913 | 0.396 | N/A |
|  | Bagging |  | 0.515 | 0.832 [0.0324] |  | 0.838 | 0.925 | 0.523 | N/A |
|  | Stacking (RF + KNN) |  | 0.748 | 0.944 [0.0189] |  | 0.938 | 0.938 | 0.765 | N/A |
| Sonia, 2023 | Deep neural network (DNN) | Cardiovascular disease | 0.9028 | 0.851 |  | 0.9585 | N/A | 0.901 | 0.851 |
|  | Inception |  | 0.83 | 0.82 |  | 0.86 | N/A | 0.82 | 0.8 |
|  | AlexNet |  | 0.92 | 0.871 |  | 0.966 | N/A | 0.918 | 0.878 |
|  | VGG-16 |  | 0.891 | 0.851 |  | 0.938 | N/A | 0.891 | 0.861 |
|  | LeNet-5 |  | 0.869 | 0.714 |  | 0.829 | N/A | 0.766 | 0.707 |
|  | LSTM |  | 0.861 | 0.909 |  | 0.863 | N/A | 0.886 | 0.908 |
|  | DNHRV |  | 0.954 | 0.959 |  | 0.989 | N/A | 0.988 | 0.941 |
| Ding, 2023 | Logistic regression | ASCVD (coronary artery disease, cerebrovascular disease, peripheral vascular disease) | 0.746 | 0.809 [0.0149] |  | 0.716 | 0.786 | 0.75 | N/A |
|  | SVM |  | 0.77 | 0.811 [0.0149] |  | 0.77 | 0.757 | 0.764 | N/A |
|  | AdaBoost |  | 0.77 | 0.834 [0.0141] |  | 0.77 | 0.814 | 0.792 | N/A |
|  | Gradient boosting decision tree (GBDT) |  | 0.824 | 0.855 [0.0133] |  | 0.824 | 0.786 | 0.813 | N/A |
|  | RF |  | 0.838 | 0.859 [0.0131] |  | 0.838 | 0.814 | 0.832 | N/A |
|  | Logistic regression | ASCVD *External validation | 0.798 | 0.8 [0.0152] |  | 0.75 | 0.711 | 0.712 | N/A |
|  | SVM |  | 0.79 | 0.812 [0.0148] |  | 0.75 | 0.7 | 0.701 | N/A |
|  | AdaBoost |  | 0.835 | 0.82 [0.0146] |  | 0.833 | 0.763 | 0.766 | N/A |
|  | Gradient boosting decision tree (GBDT) |  | 0.835 | 0.821 [0.0145] |  | 0.833 | 0.763 | 0.766 | N/A |
|  | RF |  | 0.857 | 0.823 [0.0145] |  | 0.833 | 0.797 | 0.799 | N/A |
| Abegaz, 2023 | RF | Major Adverse Cardiovascular Events (MACE) | 0.85 | 0.8 |  | 0.9 | N/A | 0.78 [0.76-0.80] | 0.81 |
|  | XGBoost |  | 0.87 | 0.82 |  | 0.93 | N/A | 0.80 [0.78-0.82] | 0.82 |
|  | LR |  | 0.7 | 0.73 |  | 0.6 | N/A | 0.65 [0.62-0.67] | 0.85 |
|  | Weighted ensemble model (WEM) |  | 0.84 | 0.75 |  | 0.9 | N/A | 0.75 [0.73-0.76] | 0.78 |
|  | RF | Myocardial Infarction (MI) | 0.91 | 0.74 |  |  | N/A | 0.85 [0.84-0.87] | 0.87 |
|  | XGBoost |  | 0.92 | 0.77 |  |  | N/A | 0.85 [0.84-0.87] | 0.86 |
|  | LR |  | 0.74 | 0.72 |  |  | N/A | 0.64 [0.62-0.66] | 0.92 |
|  | Weighted ensemble model (WEM) |  | 0.92 | 0.73 |  |  | N/A | 0.86 [0.83-0.87] | 0.87 |
|  | RF | Heart failure (HF) | 0.86 | 0.78 |  |  | N/A | 0.80 [0.78-0.82] | 0.82 |
|  | XGBoost |  | 0.89 | 0.8 |  |  | N/A | 0.81 [0.79-0.83] | 0.84 |
|  | LR |  | 0.74 | 0.73 |  |  | N/A | 0.66 [0.64-0.68] | 0.89 |
|  | Weighted ensemble model (WEM) |  | 0.86 | 0.74 |  |  | N/A | 0.76 [0.74-0.78] | 0.8 |
|  | RF | Stroke | 0.98 | 0.71 |  |  | N/A | 0.97 [0.95-0.98] | 0.96 |
|  | XGBoost |  | 0.92 | 0.74 |  |  | N/A | 0.97 [0.96-0.98] | 0.96 |
|  | LR |  | 0.77 | 0.7 |  |  | N/A | 0.63 [0.61-0.65] | 0.98 |
|  | Weighted ensemble model (WEM) |  | 0.95 | 0.74 |  |  | N/A | 0.97 [0.96-0.99] | 0.94 |
| Vimont, 2023 | LR | Cardiovascular disease (Heart failure, peripheral arterial disease, MI, stroke, UA, TIA, CV-related death) | N/A | 0.715 [0.658 - 0.712] |  | N/A | N/A | N/A | N/A |
|  | RF |  | N/A | 0.786 [0.747 - 0.825] |  | N/A | N/A | N/A | N/A |
|  | Neural network (NN) |  | N/A | 0.738 [0.686 - 0.79] |  | N/A | N/A | N/A | N/A |
| Gandin, 2023 | Cox proportional hazards regression | 5-year heart failure | N/A | 0.77 [0.732 - 0.807] |  | N/A | N/A | N/A | N/A |
|  | Non-linear proportional hazards deep neural network (PHNN) |  | N/A | 0.78 [0.743 - 0.817] |  | N/A | N/A | N/A | N/A |
| Zhong, 2022 | LR | Acute coronary syndrome | 0.73 | 0.86 |  | 0.7 | N/A | 0.8 | 0.76 |
|  | LR with lasso |  | 0.67 | 0.8 |  | 0.59 | N/A | 0.74 | 0.77 |
|  | KNN |  | 0.74 | 0.88 |  | 0.64 | N/A | 0.8 | 0.89 |
|  | SVM linear |  | 0.78 | 0.9 |  | 0.75 | N/A | 0.82 | 0.83 |
|  | SVM radial |  | **0.8** | 0.92 |  | 0.76 | N/A | 0.83 | 0.84 |
|  | Decision tree |  | 0.78 | 0.88 |  | 0.75 | N/A | 0.82 | 0.83 |
|  | Random forest |  | 0.87 | 0.96 |  | 0.83 | N/A | 0.89 | 0.91 |
|  | XGBoost |  | 0.86 | 0.96 |  | 0.81 | N/A | 0.88 | 0.91 |
|  | ANN |  | 0.52 | 0.7 |  | 0.38 | N/A | 0.68 | 0.8 |
| Momenzadeh, 2022 | SVC | Cardiovascular disease | N/A | 0.672 |  | N/A | N/A | N/A | N/A |
|  | GB |  | N/A | 0.694 |  | N/A | N/A | N/A | N/A |
|  | ET |  | N/A | 0.699 |  | N/A | N/A | N/A | N/A |
|  | RF |  | N/A | 0.693 |  | N/A | N/A | N/A | N/A |
|  | AdaBoost |  | N/A | 0.688 |  | N/A | N/A | N/A | N/A |
|  | LR |  | N/A | 0.679 |  | N/A | N/A | N/A | N/A |
| Nicolucci, 2022 | XGBoost | Cardiovascular disease | N/A | 0.817 |  | 70.5 | 75.8 | 74.8 | N/A |
|  |  | Cerebrovascular disease | N/A | 0.846 |  | 89.1 | 59.2 | 70.5 | N/A |
|  |  | Peripheral vascular disease | N/A | 0.857 |  | 72.2 | 82.1 | 80.5 | N/A |
|  |  | Cardiovascular disease* | N/A | 0.7615 |  | N/A | N/A | 48.5-82 | N/A |
|  |  | Cerebrovascular disease* | N/A | 0.725 |  | N/A | N/A | 58.5-83.9 | N/A |
|  |  | Peripheral vascular disease* | N/A | 0.7275 |  | N/A | N/A | 60.2-75.4 | N/A |
| Hong, 2021 | Cox proportional hazard  LASSO regression | Coronary heart disease  Heart failure  Stroke | N/A | 0.85 |  | N/A | N/A | N/A | N/A |
| Aminian, 2020 | Regression (Cox proportional hazards, exponential, Fine-Gray)  Random Forest (RF) | Coronary artery events | N/A | 0.66 |  | N/A | N/A | N/A | N/A |
|  |  | Heart failure | N/A | 0.73 |  | N/A | N/A | N/A | N/A |
| Athanasiou, 2020 | XGBoost | CVD | N/A | 71.13 |  | 71 | N/A | N/A | N/A |
| Zarkogianni, 2018 | Hybrid ensemble | CVD (CHD, Stroke) | N/A | 71.48 |  | 61 | 72.64 | 71.79 | N/A |
|  | BLR model |  | N/A | 55.11 |  | 67.5 | 39.08 | 41.25 | N/A |
|  | FFN |  | N/A | 60.09 |  | 57 | 55.49 | 55.54 | N/A |
|  | CART |  | N/A | 47.99 |  | 11.5 | 83.84 | 78.57 | N/A |
|  | RF |  | N/A | 60.8 |  | 37 | 70.53 | 68.04 | N/A |
|  | NB |  | N/A | 67.19 |  | 58.5 | 68.79 | 68.04 | N/A |
| Segar, 2019 | Random survival forest (RSF) | Heart failure | N/A | 0.77 |  | N/A | N/A | N/A | N/A |
|  | Cox-based method |  | N/A | 0.73 |  | N/A | N/A | N/A | N/A |
|  | Random survival forest (RSF) |  | N/A | 0.74 |  |  |  |  |  |
|  | Cox-based method |  | N/A | 0.7 |  |  |  |  |  |
| Fan, 2020 | Random forest (RF) | CV (Coronary Heart Disease) | Available but not extracted here | 0.77 |  | Available but not extracted here | Not provided | Available but not extracted here | Available but not extracted here |
|  |  | CV (Coronary Heart Disease)* | Available but not extracted here | 0.71 |  | Available but not extracted here | Not provided | Available but not extracted here | Available but not extracted here |
| Xu, 2017 | LR | Stroke |  | 0.7859 |  |  |  | 0.8548 |  |
| Lee, 2023 | GRU-ODE-Bayes-based | CVD | 0.39±0.033 | 0.812 |  | 0.251±0.027 | 0.994±0.003 |  | 0.889±0.041 |
|  | XGBoost |  | 0.364±0.047 | 0.78 |  | 0.233±0.036 | 0.993±0.004 |  | 0.842±0.065 |
| Wang, 2023 | RF | Diabetic foot | N/A | 0.808 |  | N/A | N/A | N/A | N/A |
| Kanda, 2022 | XGBoost | Heart failure diagnosis | N/A | 0.799 |  | N/A | N/A | N/A | N/A |
|  |  | Heart failure hospitalization | N/A | 0.809 |  | N/A | N/A | N/A | N/A |
|  |  | MACE | N/A | 0.792 |  | N/A | N/A | N/A | N/A |
|  |  | Heart failure diagnosis* | N/A | 0.752 |  | N/A | N/A | N/A | N/A |
|  |  | Heart failure hospitalization* | N/A | 0.898 |  | N/A | N/A | N/A | N/A |
|  |  | MACE* | N/A | 0.743 |  | N/A | N/A | N/A | N/A |
| Lee, 2021 | Conditional inference survival forest (CISF) | Acute myocardial infarction |  | 0.927 |  | 0.8851 |  |  | 0.9083 |
|  | Random survival forest (RSF) |  |  | 0.8506 |  | 0.8606 |  |  | 0.8634 |
|  | Multivariate Cox model |  | N/A | 0.7568 |  | 0.7568 |  |  | 0.8197 |
| Liu, 2020 | Single-task learning with Lasso | Vascular Disease (peripheral, cardiovascular, or cerebral) | N/A | 0.6667 |  | N/A | N/A | N/A | N/A |
|  | Multi-task feature learning (MTFL) |  | N/A | 0.707 |  | N/A | N/A | N/A | N/A |
|  | Outcome-specific multi-task learning (OS-MTL) |  | N/A | 0.7213 |  | N/A | N/A | N/A | N/A |
|  | Private-shared MTL |  | N/A | 0.7463 |  | N/A | N/A | N/A | N/A |
| Longato, 2020 | Neural network (NN) | 4P-MACE | N/A | 0.791 |  | N/A | N/A | N/A | N/A |
|  | Neural network (NN) | Heart failure | N/A | 0.796 |  | N/A | N/A | N/A | N/A |
|  | Neural network (NN) | Stroke | N/A | 0.714 |  | N/A | N/A | N/A | N/A |
|  | Neural network (NN) | Myocardial infarction | N/A | 0.708 |  | Not Provided | Not Provided | Not Provided | Not Provided |
| Phan, 2023 | Logistic Regression (LR) | Ischemic Stroke | N/A | 0.75 |  | Available but not extracted here | Available but not extracted here | Available but not extracted here | Available but not extracted here |
|  | Linear discriminant analysis (LDA) |  | N/A | 0.76 |  | Available but not extracted here | Available but not extracted here | Available but not extracted here | Available but not extracted here |
|  | Light Gradient Boosting Machine (LGBM) |  | N/A | 0.76 |  | Available but not extracted here | Available but not extracted here | Available but not extracted here | Available but not extracted here |
|  | GBM |  | N/A | 0.75 |  | Available but not extracted here | Available but not extracted here | Available but not extracted here | Available but not extracted here |
|  | RF |  | N/A | 0.66 |  | Available but not extracted here | Available but not extracted here | Available but not extracted here | Available but not extracted here |
|  | XGBoost |  | N/A | 0.72 |  | Available but not extracted here | Available but not extracted here | Available but not extracted here | Available but not extracted here |
|  | RF |  | N/A | 0.75 |  | Available but not extracted here | Available but not extracted here | Available but not extracted here | Available but not extracted here |
|  | AdaBoost |  | N/A | 0.76 |  | Available but not extracted here | Available but not extracted here | Available but not extracted here | Available but not extracted here |
|  | Voting ensemble |  | N/A | 0.75 |  | Available but not extracted here | Available but not extracted here | Available but not extracted here | Available but not extracted here |
|  | Logistic Regression (LR) | Ischemic Stroke* | N/A | 0.72 |  | Available but not extracted here | Available but not extracted here | Available but not extracted here | Available but not extracted here |
|  | Linear discriminant analysis (LDA) |  | N/A | 0.75 |  | Available but not extracted here | Available but not extracted here | Available but not extracted here | Available but not extracted here |
|  | Light Gradient Boosting Machine (LGBM) |  | N/A | 0.96 |  | Available but not extracted here | Available but not extracted here | Available but not extracted here | Available but not extracted here |
|  | GBM |  | N/A | 0.93 |  | Available but not extracted here | Available but not extracted here | Available but not extracted here | Available but not extracted here |
|  | RF |  | N/A | 0.99 |  | Available but not extracted here | Available but not extracted here | Available but not extracted here | Available but not extracted here |
|  | XGBoost |  | N/A | 0.97 |  | Available but not extracted here | Available but not extracted here | Available but not extracted here | Available but not extracted here |
|  | RF |  | N/A | 0.91 |  | Available but not extracted here | Available but not extracted here | Available but not extracted here | Available but not extracted here |
|  | AdaBoost |  | N/A | 0.88 |  | Available but not extracted here | Available but not extracted here | Available but not extracted here | Available but not extracted here |
|  | Voting ensemble |  | N/A | 0.72 |  | Available but not extracted here | Available but not extracted here | Available but not extracted here | Available but not extracted here |
| Rahman, 2018 | LR | CVD | 0.51 | 0.72 |  | Available but not extracted here | Available but not extracted here | Available but not extracted here | Available but not extracted here |
|  | SVM |  | 0 | 0.5 |  | Available but not extracted here | Available but not extracted here | Available but not extracted here | Available but not extracted here |
|  | Naive Bayes (NB) |  | 0.3 | 0.74 |  | Available but not extracted here | Available but not extracted here | Available but not extracted here | Available but not extracted here |
|  | Decision tree |  | 0.42 | 0.68 |  | Available but not extracted here | Available but not extracted here | Available but not extracted here | Available but not extracted here |
|  | Decision tree AdaBoost |  | 0.4 | 0.67 |  | Available but not extracted here | Available but not extracted here | Available but not extracted here | Available but not extracted here |
|  | Random forest |  | 0.36 | 0.61 |  | Available but not extracted here | Available but not extracted here | Available but not extracted here | Available but not extracted here |
| Liu, 2018 | Single-task learning (STL) | Vascular Disease | N/A | 0.6111 |  | N/A | N/A | N/A | N/A |
|  | Multi-task learning (MTL) | Vascular Disease* | N/A | 0.617 |  | N/A | N/A | N/A | N/A |
| Liu, 2020 | Bayesian Network (BN) | Macrovascular Complications | N/A | 0.753 |  | 0.827 | 0.563 | N/A | N/A |
|  | BN without prior information | Macrovascular Complications | N/A | 0.749 |  | 0.855 | 0.584 | N/A | N/A |
|  | NB |  | N/A | 0.723 |  | 0.861 | 0.503 | N/A | N/A |
|  | C5.0 |  | N/A | 0.726 |  | 0.804 | 0.599 | N/A | N/A |
|  | RF |  | N/A | 0.745 |  | 0.765 | 0.633 | N/A | N/A |
|  | Bayesian Network (BN) | Diabetic foot | N/A | 0.905 |  | 1 | 0.563 | N/A | N/A |
|  | BN without prior information |  | N/A | 0.788 |  | 0.833 | 0.584 | N/A | N/A |
|  | NB |  | N/A | 0.704 |  | 1 | 0.503 | N/A | N/A |
|  | C5.0 |  | N/A | 0.851 |  | 1 | 0.599 | N/A | N/A |
|  | RF |  | N/A | 0.761 |  | 0.833 | 0.633 | N/A | N/A |
| Dworzynski, 2020 | Reference logistic regression | Heart failure | N/A | 0.74 |  | N/A | N/A | N/A | 0.808-1 |
|  | Logistic ridge regression |  | N/A | 0.77 |  | N/A | N/A | N/A |  |
|  | Random forest |  | N/A | 0.77 |  | N/A | N/A | N/A |  |
|  | Gradient boosting |  | N/A | 0.8 |  | N/A | N/A | N/A |  |
|  | Reference logistic regression | Myocardial Infarction | N/A | 0.68 |  | N/A | N/A | N/A | 0.98-1 |
|  | Logistic ridge regression |  | N/A | 0.7 |  | N/A | N/A | N/A |  |
|  | Random forest |  | N/A | 0.67 |  | N/A | N/A | N/A |  |
|  | Gradient boosting |  | N/A | 0.71 |  | N/A | N/A | N/A |  |
|  | Reference logistic regression | Stroke | N/A | 0.71 |  | N/A | N/A | N/A | 0.793--0.935 |
|  | Logistic ridge regression |  | N/A | 0.72 |  | N/A | N/A | N/A |  |
|  | Random forest |  | N/A | 0.69 |  | N/A | N/A | N/A |  |
|  | Gradient boosting |  | N/A | 0.72 |  | N/A | N/A | N/A |  |
|  | Reference logistic regression | Cardiovascular disease | N/A | 0.66 |  | N/A | N/A | N/A |  |
|  | Logistic ridge regression |  | N/A | 0.68 |  | N/A | N/A | N/A |  |
|  | Random forest |  | N/A | 0.68 |  | N/A | N/A | N/A |  |
|  | Gradient boosting |  | N/A | 0.69 |  | N/A | N/A | N/A |  |
| Liu 2018 | Logistic regression/Single task learning (STL) | Vascular Disease | N/A | 0.6581 |  | N/A | N/A | N/A | N/A |
|  | Multi-task feature learning (MTFL) |  | N/A | 0.7059 |  | N/A | N/A | N/A | N/A |
|  | Multi-task relationship learning (MTRL) |  | N/A | 0.7069 |  | N/A | N/A | N/A | N/A |
|  | Feature and task relationship learning (FETR) |  | N/A | 0.729 |  | N/A | N/A | N/A | N/A |
|  | TREFLES |  | N/A | 0.7478 |  | N/A | N/A | N/A | N/A |
| Mei 2019 | Pooled cohort equations | CVD | N/A | 0.65 |  | N/A | N/A | N/A | N/A |
| Thomas 2018 | Patient network | Stroke | N/A | 0.78 |  | 0.873 | 0.447 | N/A | N/A |
|  | Patient network | Myocardial infarction | N/A | 0.743 |  | 0.799 | 0.483 | N/A | N/A |
|  | Patient network | Heart failure | N/A | 0.776 |  | 0.851 | 0.481 | N/A | N/A |
| Kim 2019 | Multi-task learning | Ischemic Heart Disease (IHD) | N/A | 0.74 |  | N/A | N/A | N/A | N/A |
|  | LASSO |  | N/A | 0.73 |  | N/A | N/A | N/A | N/A |
|  | GBM |  | N/A | 0.65 |  | N/A | N/A | N/A | N/A |
|  | Multi-task learning | Congestive Heart Failure (CHF) | N/A | 0.78 |  | N/A | N/A | N/A | N/A |
|  | LASSO |  | N/A | 0.77 |  | N/A | N/A | N/A | N/A |
|  | GBM |  | N/A | 0.74 |  | N/A | N/A | N/A | N/A |
|  | Multi-task learning | Cerebrovascular Disease (CVD) | N/A | 0.75 |  | N/A | N/A | N/A | N/A |
|  | LASSO |  | N/A | 0.74 |  | N/A | N/A | N/A | N/A |
|  | GBM |  | N/A | 0.72 |  | N/A | N/A | N/A | N/A |
|  | Multi-task learning | Peripheral Vascular Disease (PVD) | N/A | 0.75 |  | N/A | N/A | N/A | N/A |
|  | LASSO |  | N/A | 0.74 |  | N/A | N/A | N/A | N/A |
|  | GBM |  | N/A | 0.72 |  | N/A | N/A | N/A | N/A |
| Kim 2018 | Multi-task learning | IHD (Ischemic heart disease) | N/A | 0.57 |  | N/A | N/A | N/A | N/A |
|  | Multi-task learning | PVD (peripheral vascular disease) | N/A | 0.75 |  | N/A | N/A | N/A | N/A |
|  | Multi-task learning | CHF (congestive heart failure) | N/A | 0.83 |  | N/A | N/A | N/A | N/A |
|  | Multi-task learning | CVD (Cerebrovascular disease) | N/A | 0.75 |  | N/A | N/A | N/A | N/A |
|  | Lasso-penalized Cox-regression | IHD (Ischemic heart disease) | N/A | 0.57 |  | N/A | N/A | N/A | N/A |
|  | Lasso-penalized Cox-regression | PVD (peripheral vascular disease) | N/A | 0.75 |  | N/A | N/A | N/A | N/A |
|  | Lasso-penalized Cox-regression | CHF (congestive heart failure) | N/A | 0.84 |  | N/A | N/A | N/A | N/A |
|  | Lasso-penalized Cox-regression | CVD (Cerebrovascular disease) | N/A | 0.78 |  | N/A | N/A | N/A | N/A |
|  | Multi-task learning | IHD (Ischemic heart disease) | N/A | 0.53 |  | N/A | N/A | N/A | N/A |
|  | Multi-task learning | PVD (peripheral vascular disease) | N/A | 0.61 |  | N/A | N/A | N/A | N/A |
|  | Multi-task learning | CHF (congestive heart failure) | N/A | 0.73 |  | N/A | N/A | N/A | N/A |
|  | Multi-task learning | CVD (Cerebrovascular disease) | N/A | 0.64 |  | N/A | N/A | N/A | N/A |
|  | Lasso-penalized Cox-regression | IHD (Ischemic heart disease) | N/A | 0.53 |  | N/A | N/A | N/A | N/A |
|  | Lasso-penalized Cox-regression | PVD (peripheral vascular disease) | N/A | 0.61 |  | N/A | N/A | N/A | N/A |
|  | Lasso-penalized Cox-regression | CHF (congestive heart failure) | N/A | 0.74 |  | N/A | N/A | N/A | N/A |
|  | Lasso-penalized Cox-regression | CVD (Cerebrovascular disease) | N/A | 0.68 |  | N/A | N/A | N/A | N/A |

### Supplementary Material 6. Forest plot of artificial intelligence areas under the operating curve (AUROCs) in predicting cardiovascular diabetes complications

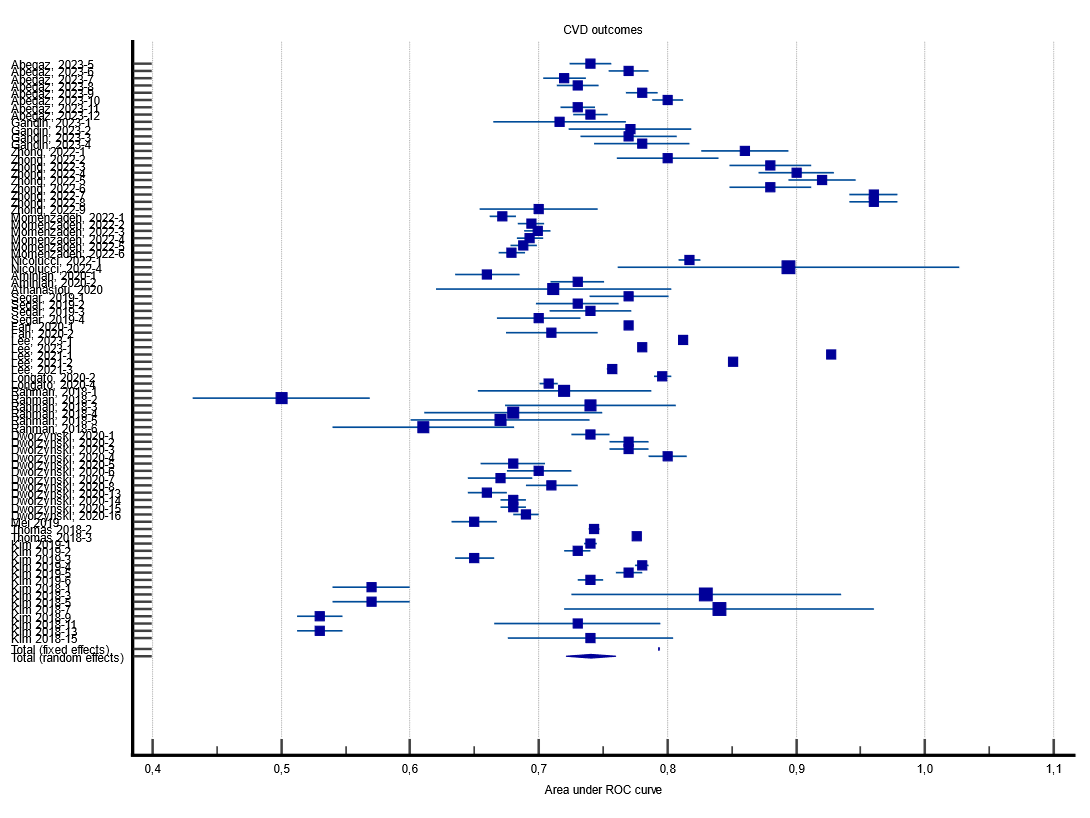

### Supplementary Material 7. Forest plot of artificial intelligence areas under the operating curve (AUROCs) in predicting peripheral vascular diabetes complications

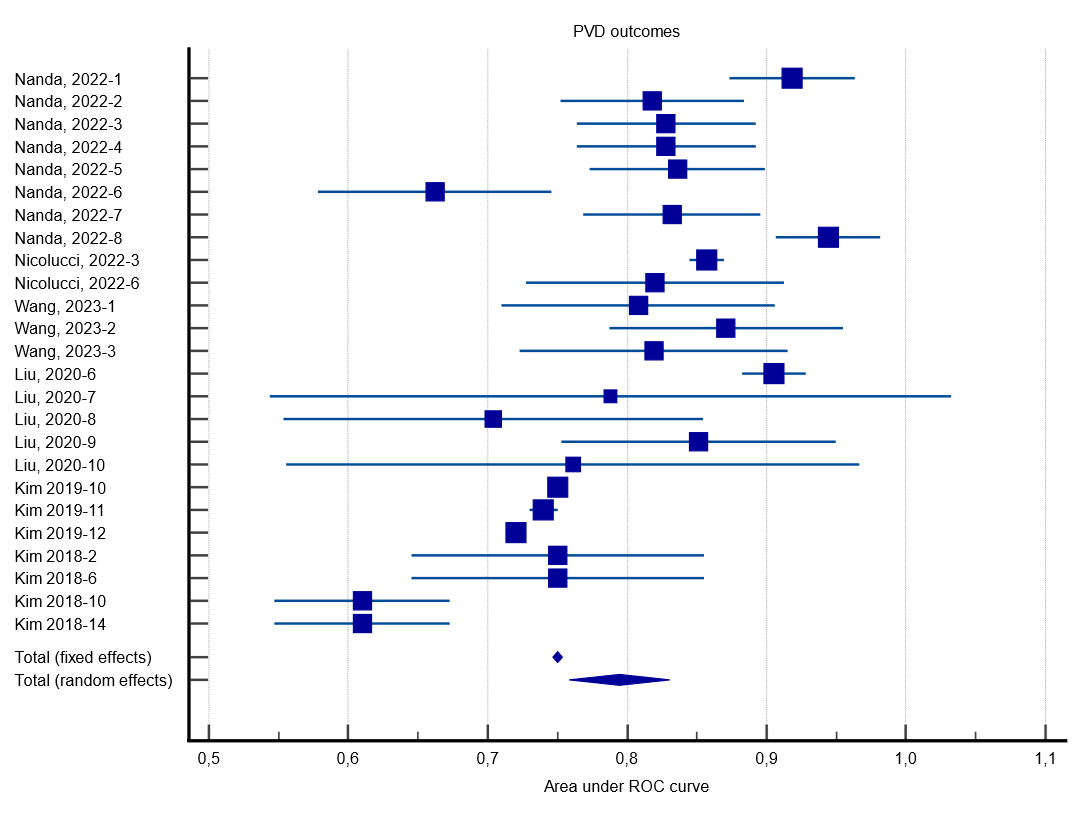

### Supplementary Material 8. Forest plot of artificial intelligence areas under the operating curve (AUROCs) in predicting cerebrovascular diabetes complications

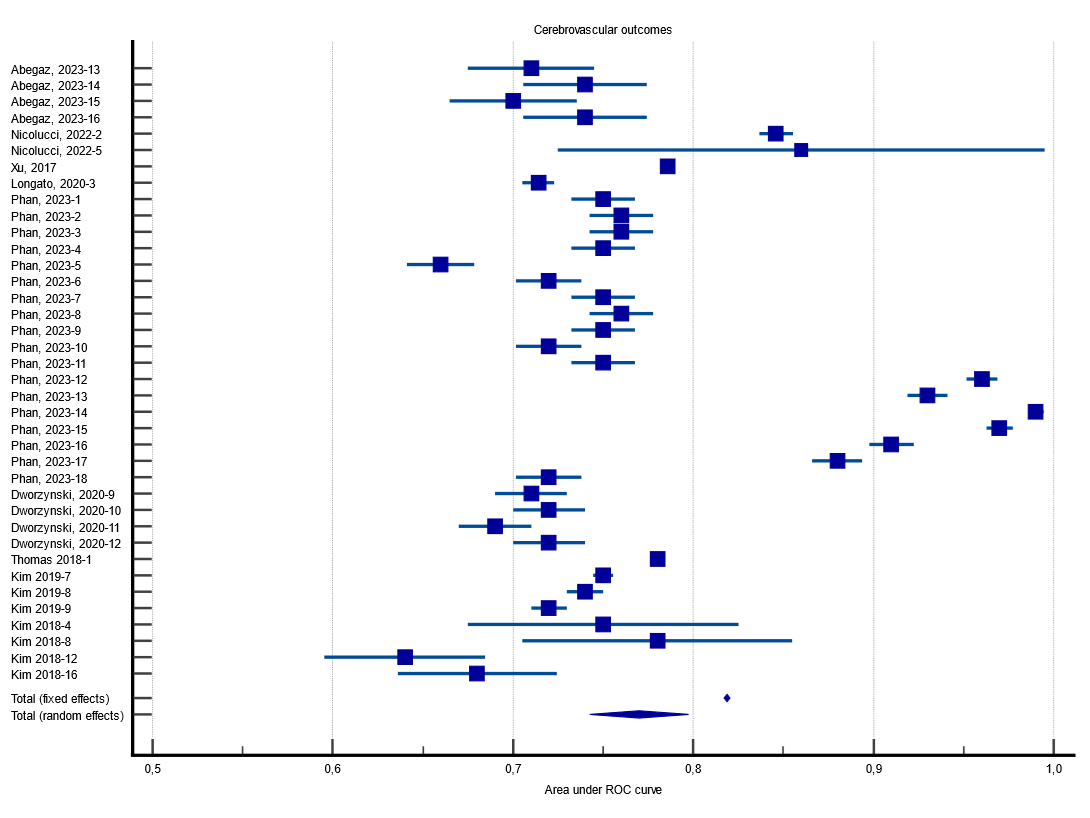

### Supplementary Material 9. Forest plot of artificial intelligence areas under the operating curve (AUROCs) in predicting mixed diabetes complications

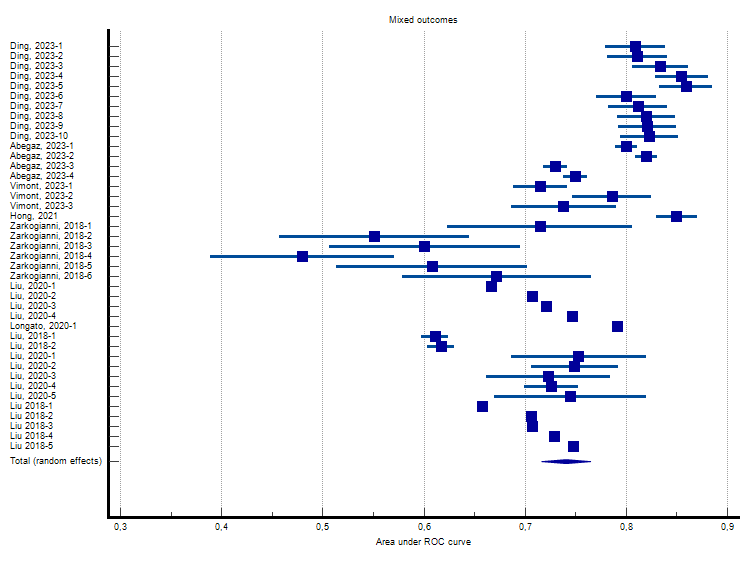

### Supplementary Material 10. Egger’s plot of artificial intelligence model performances in predicting macrovascular diabetes complications

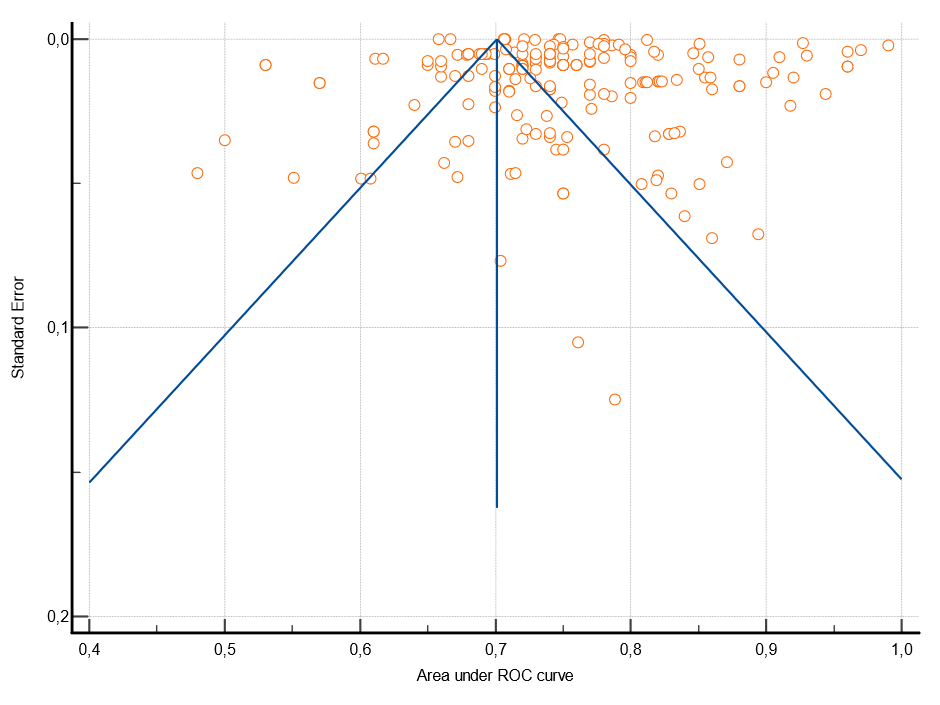

### Supplementary Material 11. Egger’s plot of artificial intelligence model performances in predicting cardiovascular diabetes complications

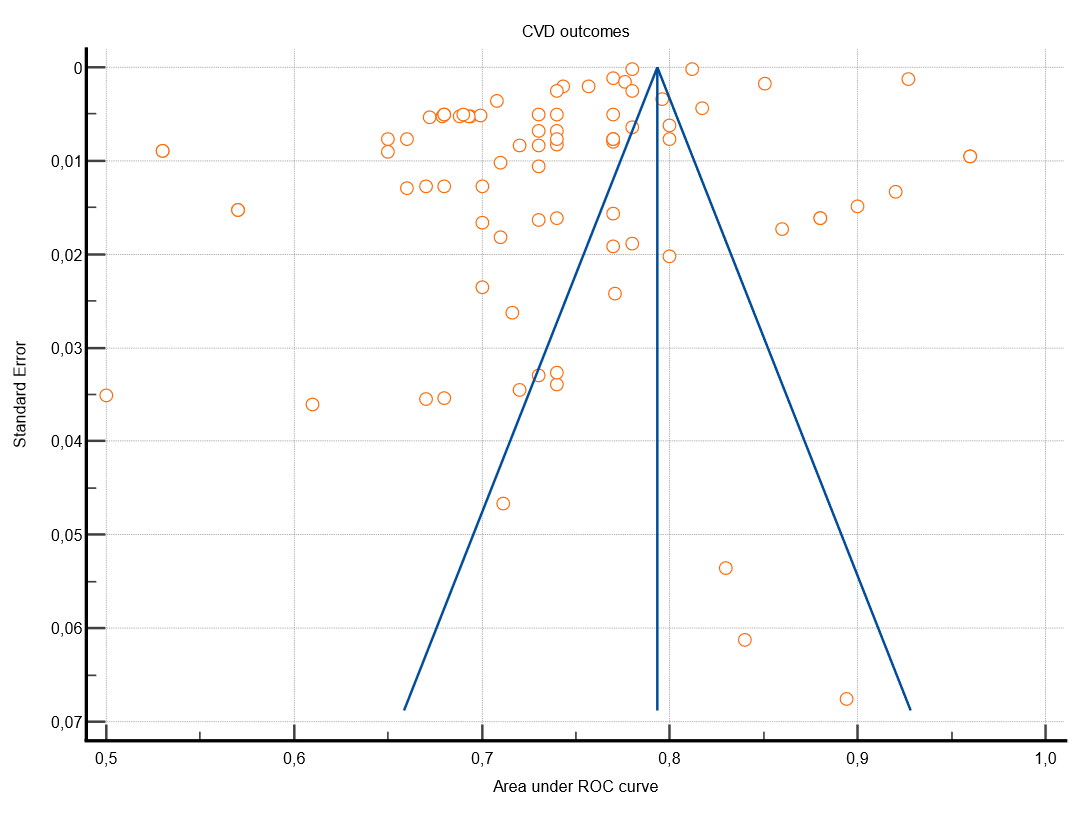

### Supplementary Material 12. Egger’s plot of artificial intelligence model performances in predicting peripheral vascular diabetes complications

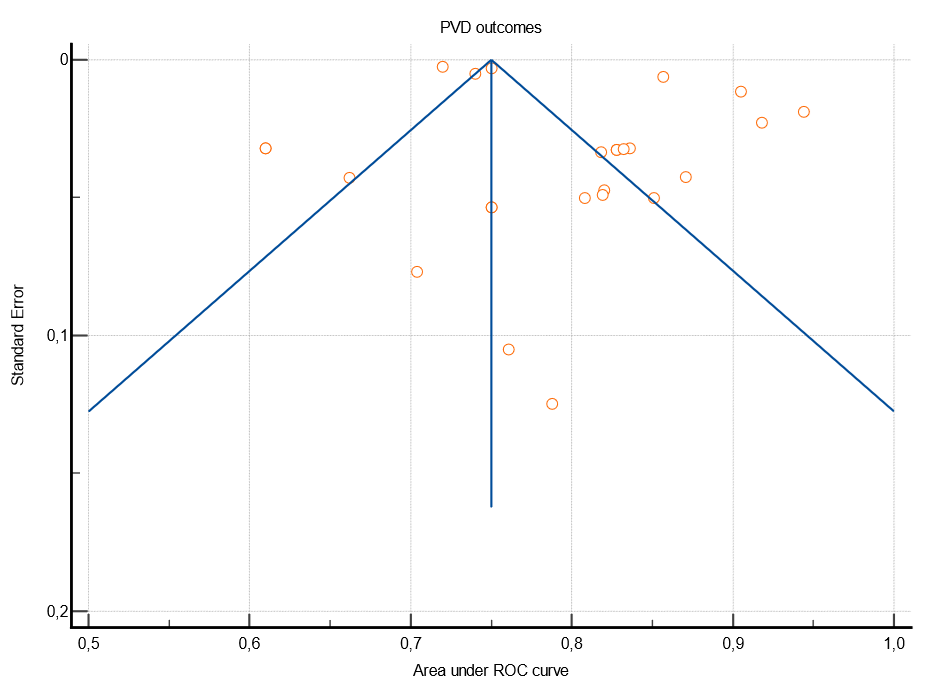

### Supplementary Material 13. Egger’s plot of artificial intelligence model performances in predicting cerebrovascular diabetes complications

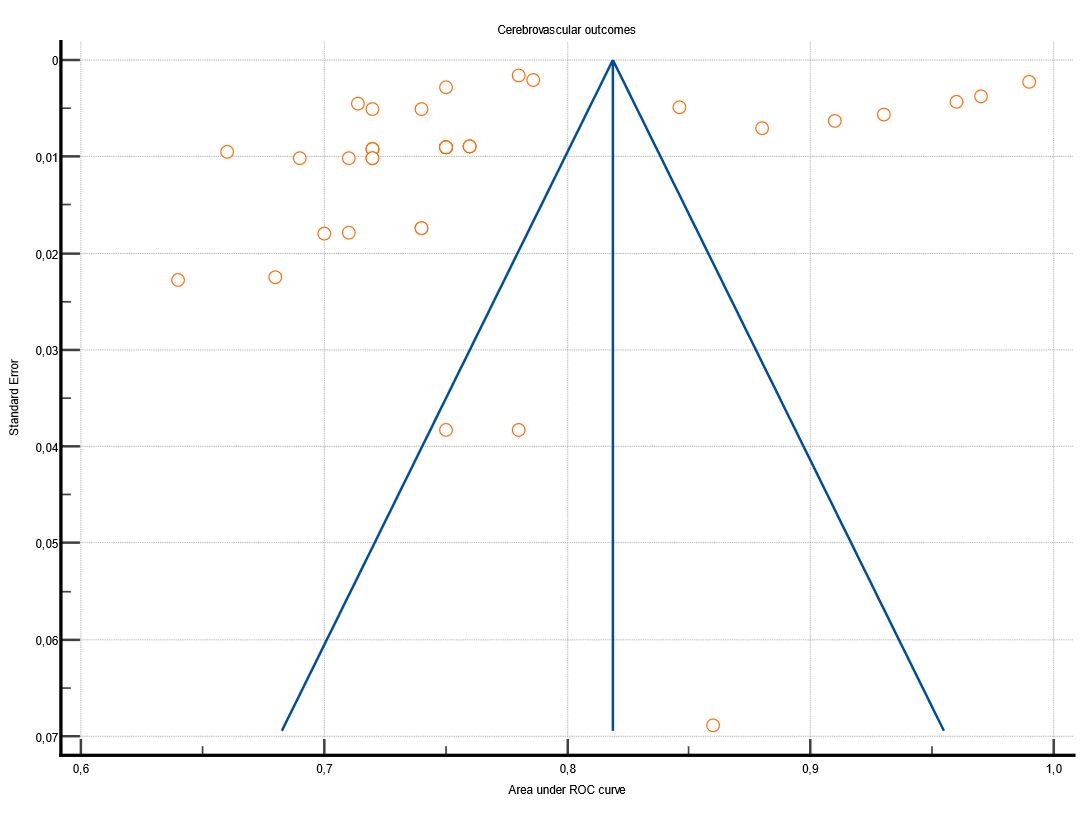

### Supplementary Material 14. Egger’s plot of artificial intelligence model performances in predicting mixed diabetes complications

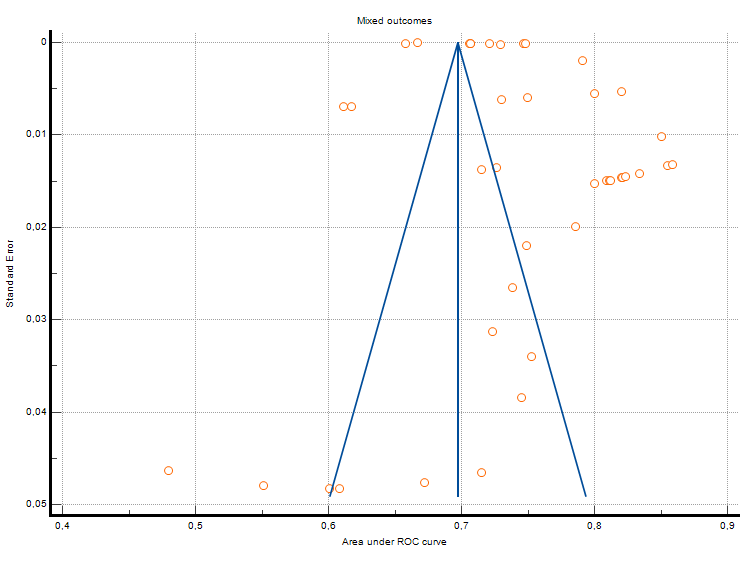
